# Cross-cohort benchmarking and validation reveals task-dependent generalisability in tuberculosis blood transcriptomic classification

**DOI:** 10.64898/2026.11.21.26368647

**Authors:** Brendan Alex, Sugam Kumar Patel, Bhavana Tiwari

## Abstract

Whole-blood transcriptomic signatures are widely proposed for tuberculosis (TB) diagnosis, but reported performance often degrades when applied to new populations. We attempt to identify how much of this can be attributed to variables like model choice, panels, tasks and cohorts. We evaluated three trained classifiers (logistic regression, random forest and a multilayer perceptron) alongside two untrained scores (singscore and directional gene average). These were applied to four published panels: Kaforou (task-specific), Zak16, Sweeney3 and Berry86. Two panels were derived internally and benchmarked: Nat(mRMR), and Nat. They were compared with 50 size-matched random panels for control and significance testing. The analysis used 952 unique samples from 10 public GEO series across three diagnostic tasks, with rotating leave-one-cohort-out (LOCO) validation. Two independent cohorts, GSE144127 active tuberculosis (ATB) vs other disease (OD) and GSE152218 (ATB vs latent tuberculosis (LTB)), served as external validation; in these, panels were compared with Holm-adjusted paired DeLong tests. Generalisability was strongly task-dependent: ATB vs LTB achieved excellent cross-cohort generalisation, consistent with previous reports. However, for ATB vs LTB random panels performed comparably, with performance only diverging when panel sizes shrank, indicating that the task itself is simpler than the others. ATB vs OD exhibited lower performance depending on the panel chosen (AUCs of 0.40-0.94; median 0.75), with the mRMR-derived panel accounting for the lower bound and failing to exceed random panels in any fold. LTB vs control (CON) showed no discernible signal across panels, models and random controls (AUC 0.39-0.78; median 0.57). Model architecture exerted minimal influence compared to the underlying biological signal, with an exception for the ATB vs OD task. Both of the final locked panels were then tested on fully independent cohorts (AUC 0.86 and 0.96). There was no significant difference between these and the published panels post Holm adjustment. An additional test on sarcoidosis had failure across all panels.

## Background

Tuberculosis (TB), caused by *Mycobacterium tuberculosis*, remains one of the leading infectious causes of mortality worldwide, with over 10 million new cases reported annually (2). An estimated one-quarter of the global population harbours latent TB infection (LTBI), most of whom remain asymptomatic; however, approximately 5–10% progress to active disease during their lifetime (3,4). These disease states are associated with distinct host immune responses that can be captured by whole-blood transcriptomic profiling (5).

Host blood transcriptomic signatures have therefore been widely explored for TB diagnosis, disease stratification, and risk prediction. For example, a 393-gene signature differentiating active TB from healthy controls was identified, their 86 gene signature similarly separates active TB infection (ATBI) from other inflammatory diseases. (5), a 16-gene signature predictive of progression from LTBI to active disease was developed (6), and a compact 3-gene signature capable of distinguishing active TB from LTBI and other diseases across multiple cohorts was derived (7).

However, inconsistent cross-cohort generalisation of whole-blood transcriptomic signatures hinders the clinical translation of machine-learning tools for diagnostics. The performance of many classifiers varies across populations, geographic settings, and study designs, reflecting biological and technical heterogeneity between cohorts. A systematic comparison of 16 published signatures revealed significant performance heterogeneity and a marked decline in discrimination upon external validation (8). Systematic validation across diverse clinical cohorts for TB prognosis was similarly shown to be necessary (9). Human populations exhibit significant variation in their transcriptomic profiles, both at rest and during disease responses (10,11). Consequently, the transcriptional shift following infection in one population frequently fails to mirror the shift in another.

The field has begun to map these generalisability challenges systematically. 72 published TB signatures were evaluated across 49 cohorts (12), while other multi-cohort frameworks have compared common machine learning architectures for TB classification (13). Despite these advances, it remains unclear how much of the variation in reported performance reflects model choice versus task definition and cohort composition, and reported failures are rarely examined for consistency in signal direction and discrimination.

In this study, we address this question using a benchmarking design in which every gene panel is scored with every model on identical training and held-out data, so that the two sources of variation can be separated and studied separately. The design uses leave-one-cohort-out (LOCO) internal cross-validation, two independent external validation cohorts; scored once, with locked models with gene selection confined to the training cohorts of each fold, size-matched random gene panels functioning as control and as a means for bootstrap confidence intervals. The individual components are established practice; what we report here is their combined application to TB across three diagnostic tasks, and the resulting observation that generalisation failure is strongly task-dependent.

## Methods

### Data

All datasets used were publicly accessible datasets published in earlier studies; these include GSE19491 (5), GSE37250 (14), and GSE73408 (15), GSE19439, GSE19444 (5), and GSE28623 (16), GSE101705 (17), GSE107994 (18), GSE42834 (19), and GSE83456 (20). Each cohort entered every task for which it had samples of both classes (ATB vs LTB: GSE19491, GSE37250, GSE73408, GSE101705, GSE107994; ATB vs OD: GSE37250, GSE73408, GSE42834, GSE83456; LTB vs CON: GSE19439, GSE19444, GSE28623, GSE107994), and all cohorts of a task took part in rotating leave-one-cohort-out validation; no discovery cohort was reserved as a fixed test set. Two further cohorts, GSE144127 (ATB vs OD) and GSE152218 (ATB vs LTB), were used only for external validation.

Several datasets were excluded for specific reasons: pediatric-focused studies were removed because host transcriptional responses to tuberculosis differ substantially between children and adults (21); datasets analysing different tissue types, such as PBMCs in GSE62525 and GSE54992, were excluded due to substantial transcriptional differences; and datasets containing comorbidities known to alter immune expression, such as HIV coinfection (5,13), were also removed. For datasets where only some samples were ineligible, samples were selected and labelled by GSM from GEO metadata and excluded for HIV coinfection, non-whole-blood tissue, extrapulmonary TB, sampling during or after treatment, submitter QC flags, or duplicate arrays. In GSE19491 and GSE22098 the OD samples (whose sub-series is confounded with OD status) and the GSE19435 ATB samples (a sub-series with no LTB comparator) were also removed to reduce risk of confound.

Note that after these exclusions, the datasets used may differ substantially from those listed on GEO. A Zenodo link contains the versions of the datasets used for the purposes of this paper. The samples used from each dataset are given in **table 1**.

**Table 1.** Samples used from each dataset (samples entering at least one task). *GSE19439 and GSE19444 are sub-series of the Superseries GSE19491.

| **Dataset** | **ATB** | **LTB** | **CON** | **OD** | **TOTAL** |
|---|---|---|---|---|---|
| GSE19491 | 54 | 69 | 0 | 0 | 123 |
| GSE37250 | 97 | 83 | 0 | 83 | 263 |
| GSE73408 | 35 | 35 | 0 | 39 | 109 |
| GSE19439* | 0 | 17 | 12 | 0 | 29 |
| GSE19444* | 0 | 21 | 12 | 0 | 33 |
| GSE28623 | 0 | 25 | 37 | 0 | 62 |
| GSE101705 | 28 | 16 | 0 | 0 | 44 |
| GSE107994 | 34 | 44 | 49 | 0 | 127 |
| GSE42834 | 35 | 0 | 0 | 71 | 106 |
| GSE83456 | 45 | 0 | 0 | 49 | 94 |
| Total | 328 | 310 | 110 | 242 | 990 |
| Total (unique) | 328 | 272 | 110 | 242 | 952 |

### Data Preprocessing

All datasets were downloaded from GEO and then converted into expression tables. Afterwards, they were pruned to exclude in treatment samples from longitudinal studies, as anti-tubercular treatment is known to alter gene expression profiles significantly, thereby complicating classification. This is in addition to the other excluded samples from earlier, which were also removed.

After these alterations to the expression matrix were applied, each cohort was processed independently using a per-gene z-score transformation, in which expression values for each gene were centred and scaled within each dataset. This harmonisation step removes dataset-specific scale and offset effects while preserving relative expression differences between samples. No samples were mixed across datasets, and no class labels were used during this normalisation; this was done to avoid the risks of inadvertent data leakage. RNA-seq data were log2-transformed before z-scoring. Genes were restricted to those measured in every cohort of a task. X- and Y-linked genes were also removed, along with uncharacterised loci: blank or missing symbols and symbols containing PHAGE or matching LOC <digits> or C<digits>ORF<digits>. All samples passing the exclusions contributed to their own cohort’s z-scoring, including samples whose class took no part in any task.

For the independent RNA-seq cohort GSE152218, counts were normalised using DESeq2 size factors (range 0.41–2.04) and then log2(x + 1)-transformed before per-cohort z-scoring (22).

Per-cohort z-scoring was preferred over cross-cohort batch-correction methods such as ComBat (23). Such methods estimate correction parameters from information pooled across all samples included in the correction step; applying them jointly to training and external cohorts would allow information from held-out data to influence the training representation, which is the form of leakage this study was designed to exclude. Applying them to each cohort independently, by contrast, offers no advantage over per-gene standardisation. Per-cohort z-scoring, therefore, preserves strict separation between training and external data, at the cost of leaving any residual platform-related variation uncorrected.

After this process was completed, the datasets were used for feature selection, model training, and evaluation, with strictly separated training and testing splits, as shown in (**Fig. 1**).

**Fig 1.**
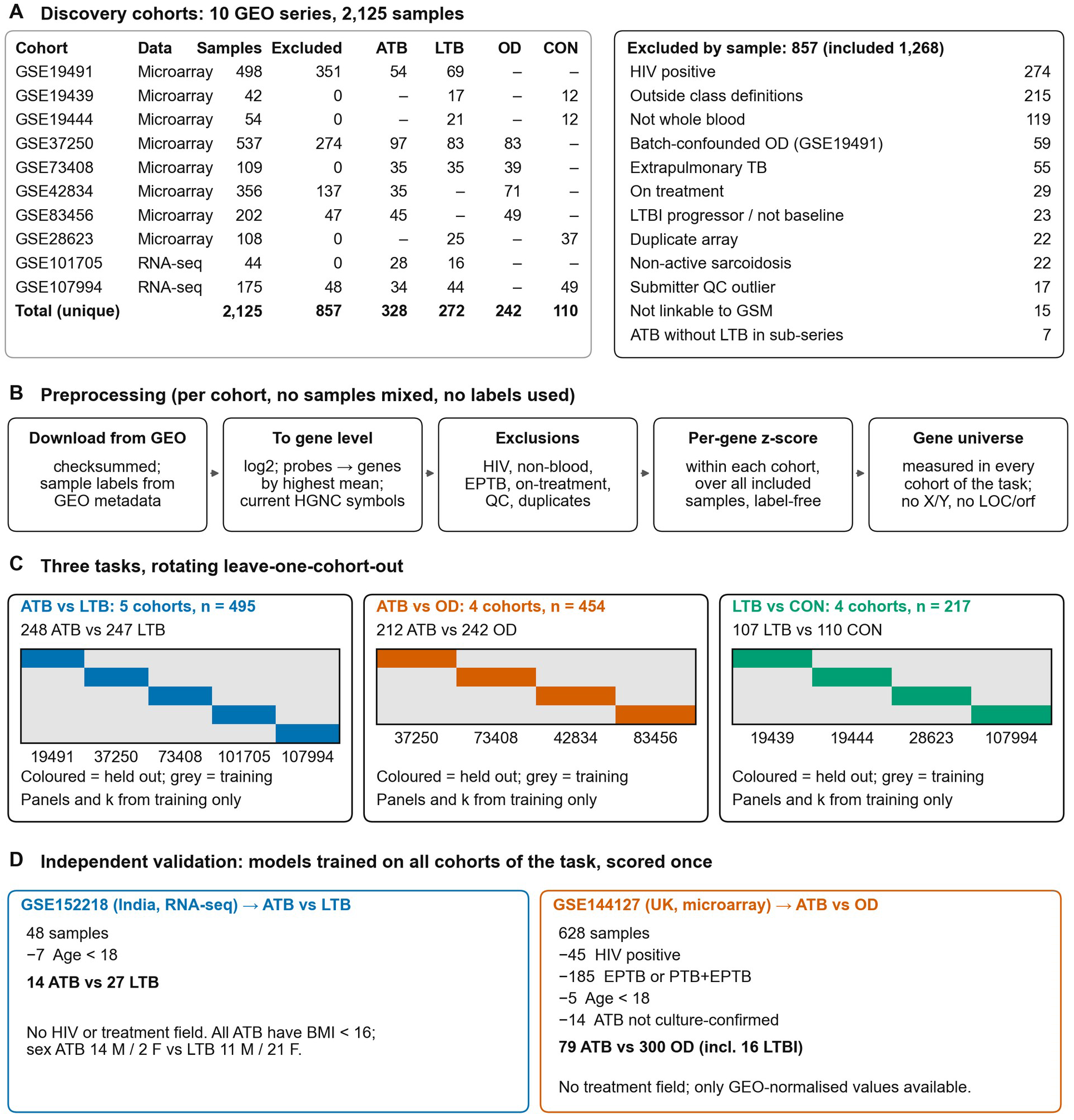
Study design, preprocessing and cohort flow. **(A)** Discovery cohorts: samples per GEO series, samples excluded, and the samples used in at least one task, by class. 2,125 samples, 857 excluded, 1,268 retained after these exclusions; of those, the 952 unique samples carrying one of the four class labels enter at least one task and are the samples counted in the class columns. Exclusions by reason (one reason per sample): HIV positive: 274 Outside class definitions: 215 Not whole blood: 119 Batch-confounded OD (GSE19491): 59 Extrapulmonary TB: 55 On treatment: 29 LTBI progressor / not baseline: 23 Duplicate array: 22 Non-active sarcoidosis: 22 Submitter QC outlier: 17 Not linkable to GSM: 15 ATB without LTB in sub-series: 7 Totals count unique samples: GSE19439 and GSE19444 are sub-series of GSE19491, so their 38 LTB samples appear in two rows but once in the total**. (B)** Preprocessing. Each cohort was processed independently; no samples were mixed across cohorts and no class labels were used at any step. **(C)** The three tasks and rotating leave-one-cohort-out validation. Each row is one fold; the coloured cohort is held out and all gene selection, model fitting and the choice of panel size use the grey training cohorts only. Cohorts are labelled by GEO accession number with the GSE prefix omitted.**(D)** Independent validation cohorts, with filters applied in the order shown: GSE144127 79 culture-confirmed pulmonary ATB vs 300 other diseases (the 16 patients with latent infection and a second diagnosis counted as OD), and GSE152218 14 ATB vs 27 LTB.

### Feature Selection

Two panels were derived within each training fold, using the training cohorts only. For Nat(mRMR), feature selection was performed separately for each task and fold using a multi-stage pipeline (24,25) as shown in (**Fig. 2**). Genes were first ranked by their mutual information with the class labels using a nonparametric k-nearest-neighbour estimator, and the top 2000 genes were retained per dataset.

**Fig 2.**
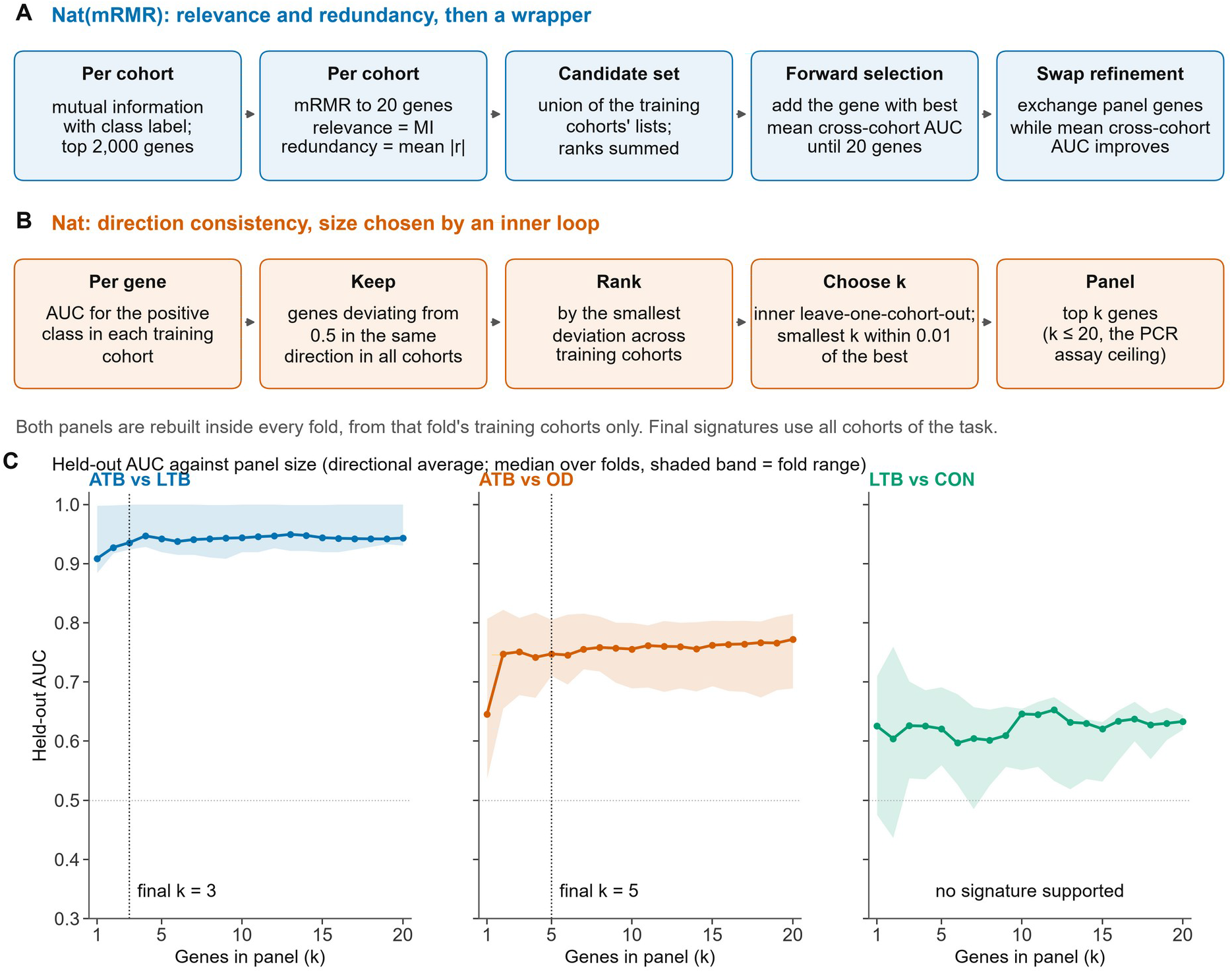
Derivation of the two internally generated panels. **(A)** Nat(mRMR), a conventional filter-and-wrapper pipeline: mutual-information filtering and a minimum-redundancy-maximum-relevance criterion within each training cohort, then forward selection and swap refinement on mean cross-cohort AUC. Relevance is mutual information (approximately 0.1-0.6 in these data) and redundancy the mean absolute correlation with the genes already selected (0-1). **(B)** Nat, selected by direction consistency: genes whose AUC for the positive class deviates from 0.5 in the same direction in every training cohort, ranked in descending order of that smallest deviation, so that the gene whose weakest training cohort separates the classes most strongly ranks first (a maximin criterion), truncated at a size k chosen by an inner leave-one-cohort-out loop over the training cohorts (smallest k within 0.01 of the best mean inner AUC, chosen separately for each scoring method). **(C)** Held-out AUC of Nat against panel size under the directional average: median across folds with the fold range shaded, and the final all-cohort k marked. Final signatures: ATB vs LTB: ANKRD22, FCGR1A, PSTPIP2 (k = 3); ATB vs OD: APOL2, WDFY1, BATF2, POLB, GBP2 (k = 5); no signature was supported for LTB vs CON.

A minimum-redundancy-maximum-relevance (mRMR) criterion was then applied to this subset to identify a compact panel of 20 genes per dataset per task, maximising relevance while minimising redundancy (24). The per-dataset mRMR top-20 gene lists were then combined by union to form the allowed set (all candidates were already restricted to genes measured in every cohort of the task). The allowed set was re-ranked by summing each training dataset’s mRMR rank, and the top five genes by aggregate rank were used as a seed for forward selection. Forward selection then iteratively added genes from the allowed universe that maximised mean cross-dataset AUC, evaluated using logistic regression trained on each dataset and tested on all others, until a 20-gene panel was reached. A swap refinement step followed, in which each panel gene was tested for replacement by any universe gene not currently in the panel; swaps were accepted if they improved mean cross-dataset AUC, and the process was repeated until no further improvement was possible.

This procedure was repeated in every fold, so Nat(mRMR) differs between folds. Nat was derived in each fold from the same training cohorts. For every gene, the AUC for the positive class was computed in each training cohort; genes whose AUC deviated from 0.5 in the same direction in every training cohort were scored by the smallest of those deviations and ranked in descending order of that score, so that the gene whose weakest training cohort separated the classes most strongly ranked first (a maximin criterion), and the top k genes formed the panel. k (1–20) was chosen by an inner leave-one-cohort-out loop over the training cohorts: for each k the ranking was rebuilt without the inner held-out cohort, and the smallest k whose mean inner AUC was within 0.01 of the best was selected, separately for each scoring method. Final signatures were derived in the same way on all cohorts of each task (directional average, the primary score): ANKRD22, FCGR1A and PSTPIP2 for ATB vs LTB (k = 3), and APOL2, WDFY1, BATF2, POLB and GBP2 for ATB vs OD (k = 5); no LTB vs CON signature was supported (S4 and S6 Tables). Twenty genes was used as the upper panel size because it is close to the limit for a PCR-based diagnostic assay (S4 Table).

All feature ranking and panel construction steps used the training cohorts of each fold only; the held-out cohort and the validation cohorts were never used at any stage of feature selection. This was checked by replacing the held-out cohort with random noise and confirming that the selected genes were unchanged (S10 Table). The published panels had predefined features from the publications and were used without modification apart from mapping symbols to current HGNC names.

X- and Y-linked genes were excluded from selection because ATB was more often male than the comparison class in most cohorts; when they were allowed, sex-chromosome genes entered the Nat(mRMR) panel in 9 of 13 folds. Gene symbols are reported using current HGNC-approved nomenclature. Published panels were evaluated in every cohort of their task, including cohorts that overlap their own derivation; those cells are flagged as optimistic and reported separately. All panels are reported in the supplementary material.

### Model Parameters

For the model comparison, the following setup was implemented. Machine learning approaches have been previously reviewed in the context of genomics (26). Every panel was scored with every method, so no single model is privileged in the panel comparison. The two untrained scores fit no weights: the directional average (DirAvg) is the mean z-score of the panel genes after multiplying each gene by its direction of change (+1 or −1, estimated from the pooled training cohorts), and singscore is a rank-based single-sample score using the same up/down split (27). A fixed seed of 42 was used for all stochastic components.

Logistic regression classifiers were implemented using scikit-learn with the lbfgs solver and L2 regularisation (C = 1.0), and the maximum number of iterations was set to 1,000 to ensure convergence. No class weighting was applied. All other hyperparameters were left at default values.

A multilayer perceptron (MLP) classifier was implemented using scikit-learn (28). The network consisted of a fully connected feed-forward architecture with a single hidden layer of 50 neurons and ReLU activation (29). Model optimisation was performed using the Adam optimiser with L2 regularisation (α = 10). Training was run for up to 5,000 iterations with a fixed random seed to ensure reproducibility. All other parameters were left at default values. A single hidden layer with 50 neurons was chosen to balance representational capacity and the risk of overfitting, given the modest sample size relative to the feature dimensionality.

The Random Forest classifiers were trained using scikit-learn with 500 decision trees. A tree count of 500 was selected to ensure stable variance reduction in the ensemble, as smaller forests can yield unstable estimates in high-dimensional transcriptomic settings. Trees were grown without an explicit maximum depth using the Gini impurity criterion (30). Bootstrap sampling was enabled, and a fixed random seed was used to ensure reproducibility. All other hyperparameters were left at default values.

### Evaluation framework

Performance was assessed by rotating leave-one-cohort-out (LOCO) validation within each task. Each GEO series was treated as an independent cohort; in each fold one cohort was held out in its entirety, panels were derived and models trained on the remaining cohorts of the task, and the held-out cohort was scored once. Every cohort served once as the held-out cohort (13 folds: 5 for ATB vs LTB and 4 each for ATB vs OD and LTB vs CON). Results are reported per fold; predictions were not pooled across folds, and no mean across folds was computed.

External validation used two cohorts that played no part in panel derivation, model training or any earlier analysis: GSE144127 (ATB vs OD; 79 culture-confirmed pulmonary ATB vs 300 OD, after excluding HIV-positive, extrapulmonary and paediatric patients) (31) and GSE152218 (ATB vs LTB; 14 ATB vs 27 LTB adults) (32). Their independence from the discovery cohorts was checked through GEO relations, shared GSMs and sample-level expression correlation: no sample identifier was shared with any discovery cohort, and the median correlation between each validation cohort and each discovery cohort fell below the median correlation measured within a single discovery cohort. Models were trained on all cohorts of the task, with Nat at its final k, and scored once; nothing was selected or tuned on validation data. No external cohort was available for LTB vs CON.

Uncertainty is quantified within each held-out or validation cohort by non-parametric bootstrap resampling.

Kaforou was evaluated in its task-specific versions for ATB vs LTB and ATB vs OD only; Zak16, Sweeney3 and Berry86 were evaluated in all three tasks. As a reference distribution, 50 random gene panels of the same size were drawn from the task’s gene universe for every fold and panel size (fixed seeds) and scored identically by every method; the draw depends on task, held-out cohort, panel size and replicate only, so all scoring methods are compared against the same random panels; a panel was considered to beat random when at most 5% of the random panels reached its AUC. Cells in which a published panel’s derivation included the held-out cohort or one of its sub-series were flagged as optimistic, and the GSE19439 and GSE19444 folds were each flagged because the training cohorts of one included the sister sub-series of the other.

Confidence intervals were estimated using non-parametric bootstrapping of test-set predictions. For each model-panel-task combination, predicted probabilities and true labels from the held-out or validation cohort were resampled with replacement 1,000 times (seed 42). Performance metrics were recalculated for each bootstrap replicate, and the 2.5th and 97.5th percentiles of the resulting distributions were reported as 95% confidence intervals. Models were trained once per evaluation and were not retrained during bootstrap resampling.

### Statistical Comparison of Classifier Performance

In the two validation cohorts, panels were compared with paired DeLong tests of the area under the receiver operating characteristic curve (AUC), computed on identical samples (33).

For each validation cohort and scoring method, Nat was compared with every published panel using the paired DeLong test, which accounts for the correlation between AUCs measured on the same samples. Differences are reported as ΔAUC (Nat minus comparator) with 95% confidence intervals, and p-values were adjusted for multiple comparisons within each cohort and method using the Holm procedure (1), the family being the six published-panel comparators tested in that cohort and method. Within the LOCO benchmark, panels were compared only with their size-matched random distributions, without adjustment for multiplicity.

## Results

### P1: Cross-cohort generalisation across tasks

Cross Cohort performance varied dramatically across the three classification tasks, revealing task-dependent generalisability patterns (Fig. 3).

**Fig 3.**
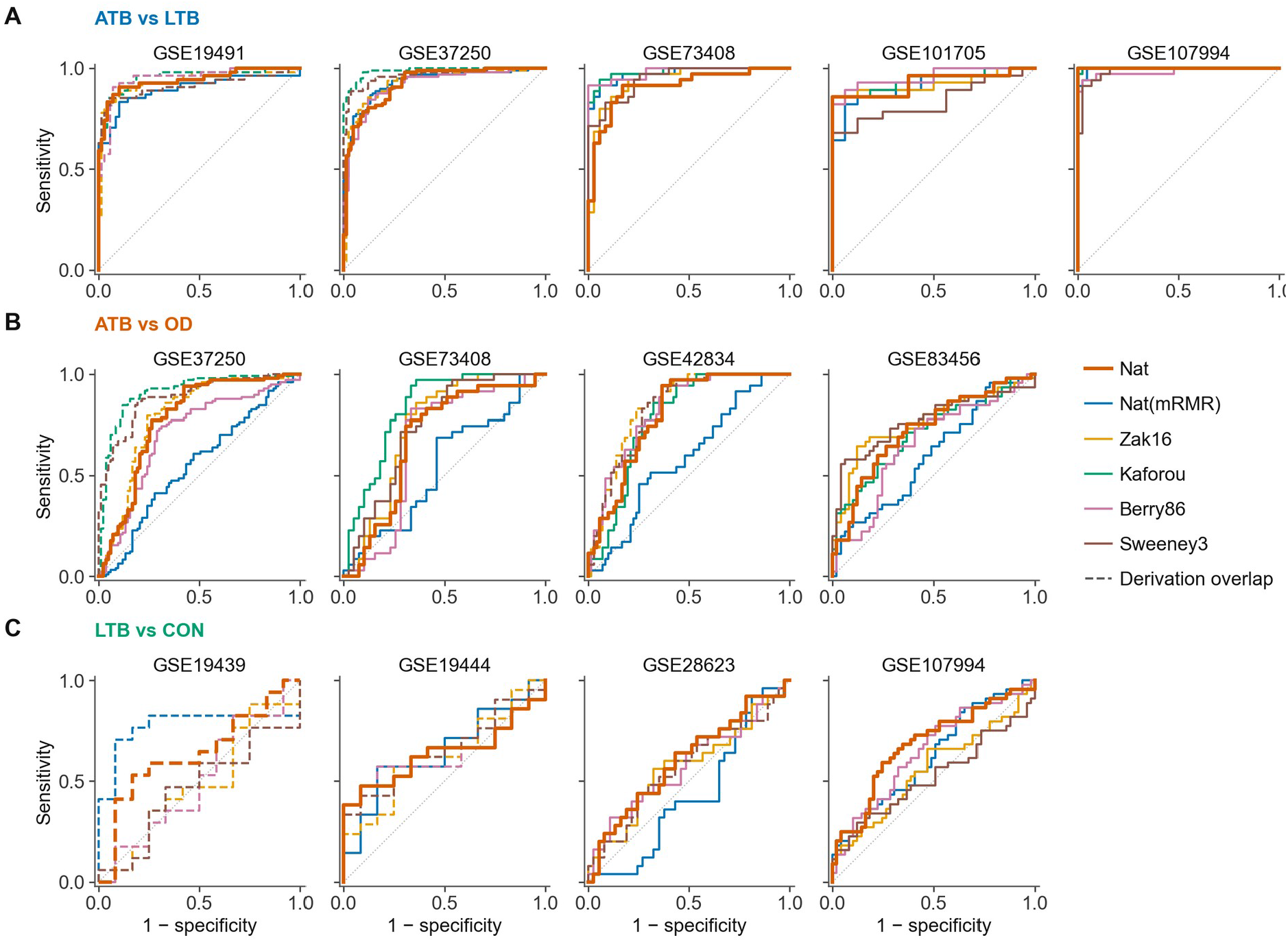
Receiver operating characteristic curves in every held-out cohort (directional average). Each panel is one-fold: the cohort named above it was withheld in full, panels were derived and models trained on the remaining cohorts of that task, and the withheld cohort was scored once. Curves are not pooled across folds. Nat is drawn heavier. Dashed curves mark cells in which the panel’s own derivation included the held-out cohort or one of its sub-series, or, for the GSE19439 fold, in which the training cohorts included the sister sub-series GSE19444; these are optimistic and are not counted in the summaries below. **(A)** ATB vs LTB, **(B)** ATB vs OD, **(C)** LTB vs CON. Kaforou is task-specific and is not evaluated for LTB vs CON. Area under the curve by fold (“*” = at most 5% of 50 size-matched random gene panels reached that value; “(d)” = derivation overlap): ATB vs LTB, GSE19491: Nat 0.941*; Nat(mRMR) 0.897*; Zak16 0.935 (d); Kaforou 0.955 (d); Berry86 0.947 (d); Sweeney3 0.914 (d) ATB vs LTB, GSE37250: Nat 0.926*; Nat(mRMR) 0.930*; Zak16 0.931 (d); Kaforou 0.987 (d); Berry86 0.917*; Sweeney3 0.967 (d) ATB vs LTB, GSE73408: Nat 0.909*; Nat(mRMR) 0.974*; Zak16 0.939; Kaforou 0.981*; Berry86 0.980*; Sweeney3 0.947* ATB vs LTB, GSE101705: Nat 0.929; Nat(mRMR) 0.915; Zak16 0.922; Kaforou 0.938; Berry86 0.955*; Sweeney3 0.837 ATB vs LTB, GSE107994: Nat 1.000*; Nat(mRMR) 0.997*; Zak16 0.999*; Kaforou 0.999*; Berry86 0.982; Sweeney3 0.985* ATB vs OD, GSE37250: Nat 0.775*; Nat(mRMR) 0.551; Zak16 0.792 (d); Kaforou 0.910 (d); Berry86 0.704*; Sweeney3 0.884 (d) ATB vs OD, GSE73408: Nat 0.684; Nat(mRMR) 0.544; Zak16 0.741*; Kaforou 0.832*; Berry86 0.661; Sweeney3 0.739* ATB vs OD, GSE42834: Nat 0.800*; Nat(mRMR) 0.584; Zak16 0.842 (d); Kaforou 0.784*; Berry86 0.826*; Sweeney3 0.830 (d) ATB vs OD, GSE83456: Nat 0.725; Nat(mRMR) 0.599; Zak16 0.761*; Kaforou 0.707*; Berry86 0.653*; Sweeney3 0.769* LTB vs CON, GSE19439: Nat 0.637 (d); Nat(mRMR) 0.775 (d); Zak16 0.485 (d); Berry86 0.505 (d); Sweeney3 0.466 (d) LTB vs CON, GSE19444: Nat 0.639; Nat(mRMR) 0.651; Zak16 0.627 (d); Berry86 0.627 (d); Sweeney3 0.655 (d) LTB vs CON, GSE28623: Nat 0.602; Nat(mRMR) 0.432; Zak16 0.572; Berry86 0.581; Sweeney3 0.572 (d) LTB vs CON, GSE107994: Nat 0.681*; Nat(mRMR) 0.624; Zak16 0.558; Berry86 0.647; Sweeney3 0.520 Clean cells beating size-matched random panels, by panel: ATB vs LTB Berry86 3/4; ATB vs LTB Kaforou 2/3; ATB vs LTB Nat 4/5; ATB vs LTB Nat(mRMR) 4/5; ATB vs LTB Sweeney3 2/3; ATB vs LTB Zak16 1/3; ATB vs OD Berry86 3/4; ATB vs OD Kaforou 3/3; ATB vs OD Nat 2/4; ATB vs OD Nat(mRMR) 0/4; ATB vs OD Sweeney3 2/2; ATB vs OD Zak16 2/2; LTB vs CON Berry86 0/2; LTB vs CON Nat 1/3; LTB vs CON Nat(mRMR) 0/3; LTB vs CON Sweeney3 0/1; LTB vs CON Zak16 0/2. Berry86 is used as published and contains the X-linked gene TLR7, which our own selection would have excluded; removing it changed held-out AUC by at most 0.024 under logistic regression, singscore and the directional average.

### ATB vs LTB Classification

This task demonstrated excellent cross-cohort generalisation. Held-out AUC was strong across models and panels (0.84-1.00; median 0.95) and consistent across folds. All five scoring approaches performed comparably (S1 Table; median held-out AUC: MLP 0.958, singscore 0.954, directional average 0.952, random forest 0.948, logistic regression 0.946).

Published gene signatures also generalised excellently. Kaforou achieved the highest median held-out AUC (0.971), followed by Berry86 (0.955), Sweeney3 (0.954) and Zak16 (0.937). Size-matched random panels also performed well, reaching median AUCs between 0.62 and 0.94 depending on panel size, indicating that active TB is associated with widespread transcriptional changes detectable by diverse gene sets.

Against these size-matched references, Nat exceeded its random comparators in 20 of 25 fold-by-method (cells) and Nat(mRMR) in 14 of 25 (**Fig. S1, Fig. S4**). The advantage over random selection depended strongly on panel size: +0.191 for Sweeney3 at three genes and +0.188 for Nat at a median of four, against +0.068 for Zak16 at 16 genes, +0.078 for Kaforou at 22, +0.044 for Nat(mRMR) at 20 and +0.028 for Berry86 at 73 showed that the random panels for this task converged with handpicked panels with increasing panel size (**Fig. 4**).

**Fig 4.**
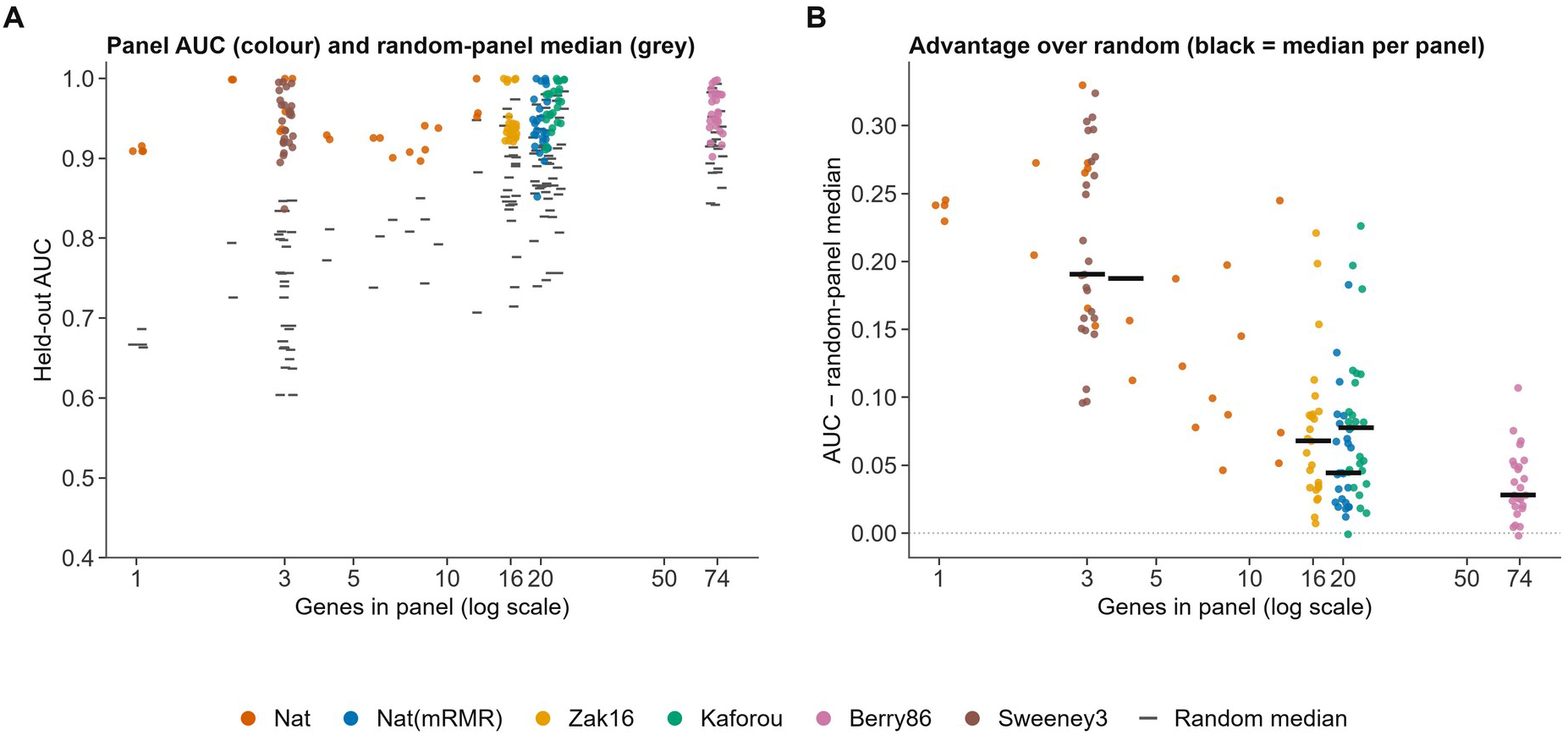
Panel size and the advantage over random gene sets, ATB vs LTB. All 150 cells (5 folds x 6 panels x 5 scoring methods). **(A)** Held-out AUC of each panel (colour) and the median AUC of the 50 size-matched random panels drawn for that fold and size (grey), against the number of genes used (log scale, small horizontal jitter). **(B)** Panel AUC minus the random-panel median; the black bar is the median per panel, drawn at its median size. Nat is plotted at the size chosen in each fold and for each method (range 1-13 genes). Median advantage over size-matched random panels (10th-90th percentile): Sweeney3 (3 genes): +0.191 (+0.122 to +0.301) Nat (4 genes): +0.188 (+0.076 to +0.271) Zak16 (16 genes): +0.068 (+0.025 to +0.138) Nat(mRMR) (20 genes): +0.044 (+0.019 to +0.102) Kaforou (22 genes): +0.078 (+0.022 to +0.156) Berry86 (73 genes): +0.028 (+0.005 to +0.067) Random panels improve steeply with size in this task, so the advantage of a curated panel falls as the panel grows.

Median performance differed by at most 0.006 between the five scoring approaches, indicating convergence of linear, tree-based, and nonlinear classifiers under strong biological separation.

### ATB vs OD Classification

Compared to ATB vs LTB classification this task had worse generalisation when testing cross cohort however the signal persists and the AUCs are much greater than 0.5. Held-out AUC ranged from 0.40 to 0.94 across models and panels (median 0.75; S2 Table). Published signatures showed variable external performance. Kaforou ATB vs OD achieved the best median AUC among curated panels (0.828), followed by Sweeney3 (0.806), Zak16 (0.758) and Berry86 (0.752). Size-matched random panels performed near chance (medians 0.51–0.61), substantially worse than most curated panels, confirming that genuine task-specific signal exists and transfers in this task.

The two internally derived panels diverged sharply. Nat exceeded its random comparators in 15/20 cells (median AUC 0.768), whereas Nat(mRMR) reached a median of 0.549 and exceeded random in 0/20 cells (**Fig. S2, Fig. S5**). The model architecture had an impact on performance here unlike the previous task, with median AUCs between 0.734 for singscore and 0.782 for the MLP. For this task the widest spread between accuracies by method is 0.048 while for ATB vs LTB it was 0.006 (Fig. 5).

**Fig 5.**
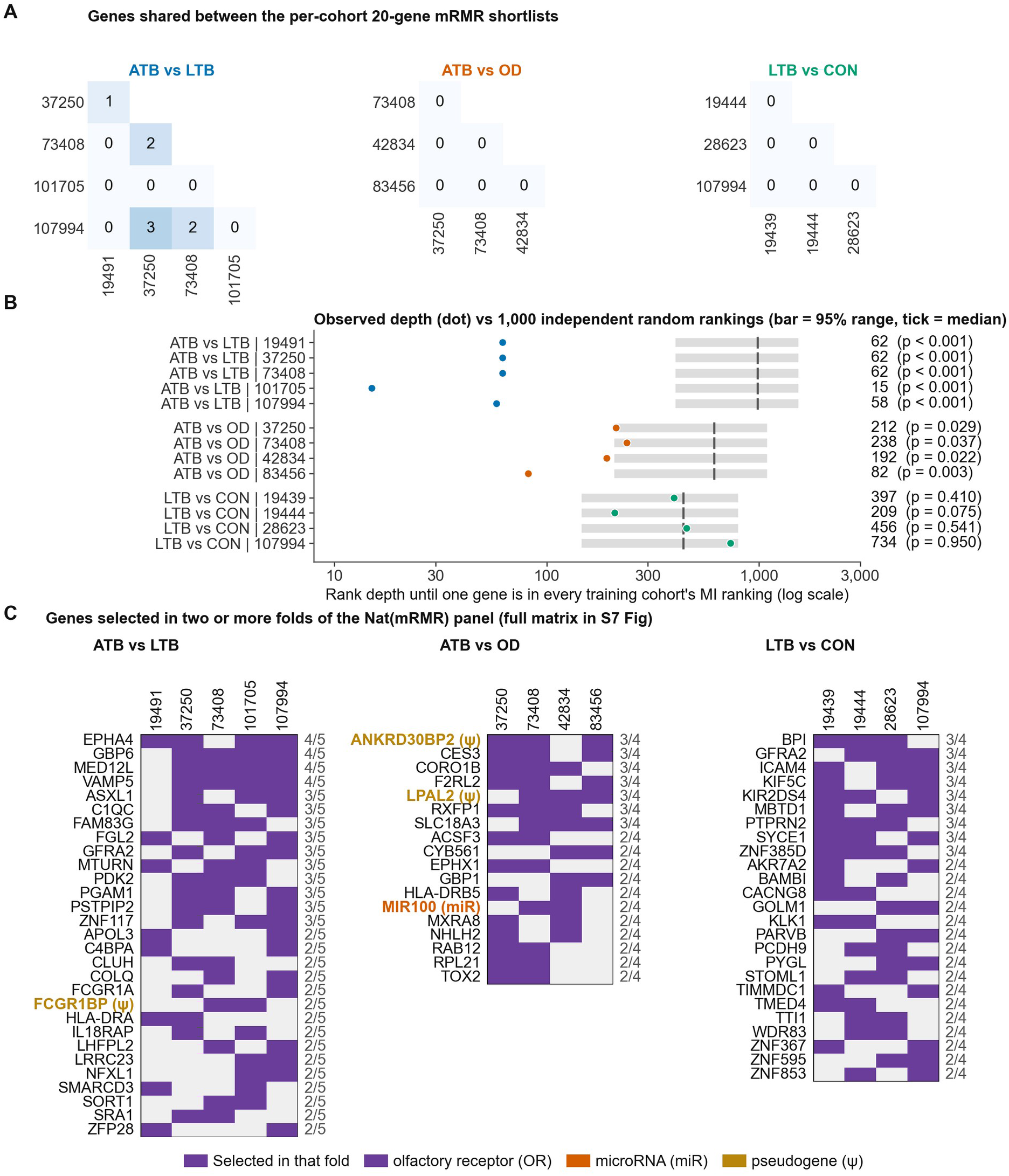
The mRMR-derived panels do not agree between cohorts and are unstable between folds. Cohorts are labelled by GEO accession number with the GSE prefix omitted. **(A)** Genes shared between the per-cohort 20-gene shortlists (mutual-information top 2,000 followed by mRMR) for every pair of cohorts in each task; shortlists are computed per cohort and do not depend on the fold. Pairs sharing any gene: ATB vs LTB 4/10 (range 0-3); ATB vs OD 0/6 (range 0-0); LTB vs CON 0/6 (range 0-0). **(B)** For each fold, the rank depth at which one gene first appears in every training cohort’s mutual-information ranking (dot) against 1,000 independent random rankings of the same gene universe (grey bar = 95% range, tick = median); p is the share of random rankings at or below the observed depth. ATB vs LTB: 15-62 vs random median 984 (p 0.000-0.000); ATB vs OD: 82-238 vs random median 615 (p 0.003-0.037); LTB vs CON: 209-734 vs random median 439 (p 0.075-0.950). **(C)** Genes selected in two or more folds of the Nat(mRMR) panel, by held-out cohort; the number at right is the count of folds selecting that gene, and coloured names are HGNC-flagged entries. No gene was selected in every fold of any task; across folds the three tasks drew on ATB vs LTB 53; ATB vs OD 55; LTB vs CON 46 distinct genes, with pairwise fold overlap of ATB vs LTB 2–11; ATB vs OD 3–10; LTB vs CON 4–8 genes. The full matrices are in **S7 Fig**.

ATB vs OD thus in comparison to ATB vs LTB is much more dependent on model choice in terms of obtaining favourable AUCs in predictions, at least in the initial cross-cohort tests, since it also has the largest gap between internal samples AUC and held out cohort tests sitting at +0.058 against −0.013 for singscore and +0.014 for DirAvg, as such while MLP does give the best AUC here it trades transfer for training fit in this task (Fig. 6).

**Fig 6.**
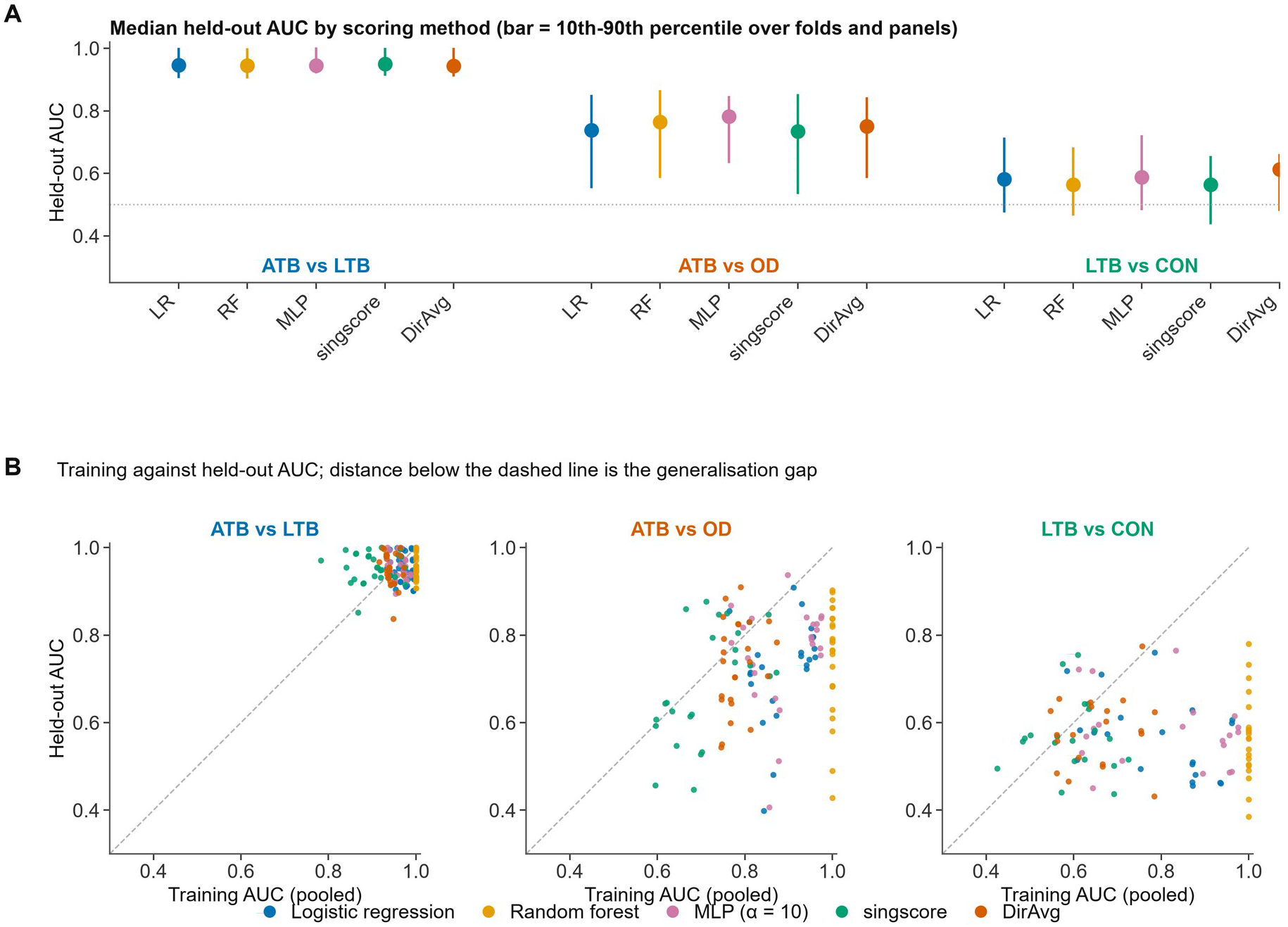
Scoring method matters less than task, and fitting buys training fit rather than transfer. **(A)** Median held-out AUC for each scoring method within each task, over all folds and panels, with the 10th-90th percentile range. Medians span 0.944-0.950 in ATB vs LTB, 0.734-0.782 in ATB vs OD and 0.563-0.613 in LTB vs CON, so the difference between methods within a task (at most 0.05) is small beside the difference between tasks (about 0.39). **(B)** Pooled training-cohort resubstitution AUC against held-out AUC for every fold, panel and method; the distance below the dashed diagonal is the generalisation gap. Median gaps across all tasks: Logistic regression +0.071, Random forest +0.169, MLP +0.058, singscore −0.013, DirAvg +0.014. The two scores that fit no weights sit closest to the diagonal; random forest is furthest below it, reaching a median gap of +0.436 in LTB vs CON.

### LTB vs CON Classification

Discrimination for this task was very poor with derived panels only slightly above chance (held-out AUC 0.39–0.78; median 0.57; S3 Table), with several cells falling at or below chance levels. Published signatures performed poorly too: Zak16 achieved the lowest median AUC (0.538), with Berry86 (0.566) and Sweeney3 (0.570) little better.

None of the curated panels evaluated here was derived for the LTB vs CON task: Zak16 was developed to predict progression from latent infection to active disease, and the remaining published panels were developed to discriminate active disease from other states. Their evaluation on this task is therefore deliberately off-label, and the results describe how these signatures behave when applied outside their intended use rather than a failure on the tasks for which they were designed. No independent validation cohort was available for this task.

No panel exceeded its size-matched random comparators in any clean fold under the directional average, with the only exceptions being Nat in the GSE107994 fold (AUC 0.681) and even that outlier performance isn’t maintained throughout and cannot be used for therapeutic purposes realistically speaking; curated panels fell mostly within or below the random-panel envelope bar the singular exception mentioned in the next paragraph. (**Fig. S3, Fig. S6**).

The five cells in which Nat(mRMR) appeared to beat random all fell in the GSE19439 fold, whose training cohorts include its sister sub-series GSE19444. Inversely the four hits from Sweeney3 were in the GSE 19444 which was the sister sub series of GSE 19439 in the training cohort. As such these two results must be viewed with some scepticism.

Median performance differed by 0.05 between scoring approaches, indicating that increased model complexity did not rescue discrimination in the absence of a stable transcriptional signal. From our testing this tasks signals seem weak if any and building a classifier seems unlikely from at least the approaches we have tried.

### P2: External testing results

#### ATB vs LTB external testing

This task demonstrated excellent performance in external tests. Held-out AUC was strong across models and panels with GSE152218 Nat DirAvg AUC at (0.963) being the best native result. The other four scoring approaches also performed very well with held-out AUCs being MLP (0.960), singscore (0.955), random forest (0.927), logistic regression (0.963) (Fig. 7B; S7 Table).

**Fig 7.**
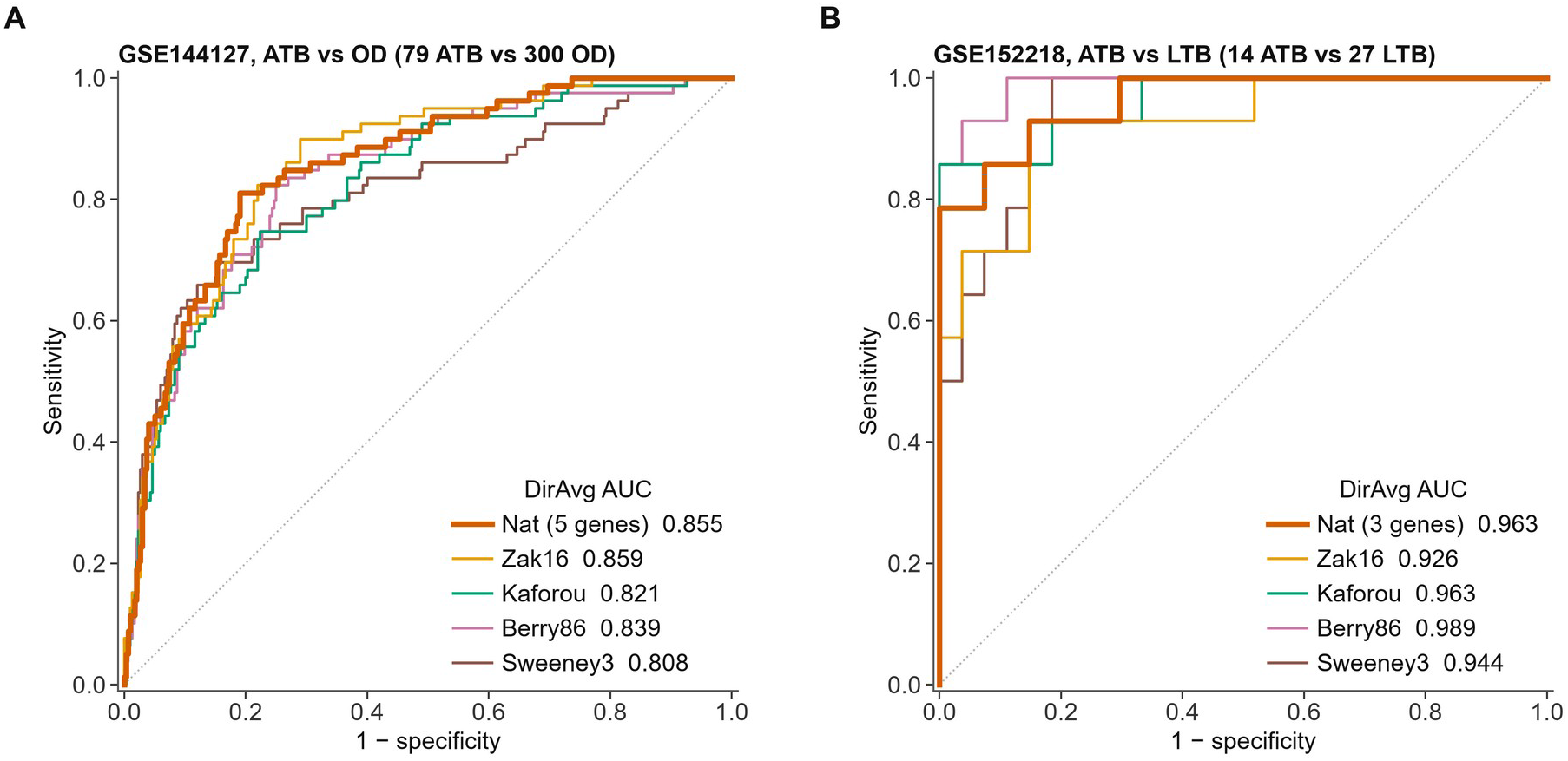
External validation of the Nat signatures in two independent cohorts. Locked models: each was trained on all cohorts of its task and scored once on the validation cohort; nothing was selected, tuned or thresholded using validation data. Curves are the directional average. Nat is APOL2, WDFY1, BATF2, POLB, GBP2 in (A) and ANKRD22, FCGR1A, PSTPIP2 in (B); areas under the curve are given in the legends. **(A)** GSE144127, 79 culture-confirmed pulmonary ATB vs 300 other diseases: Nat AUC 0.855 (95% CI 0.806-0.898). **(B)** GSE152218, 14 ATB vs 27 LTB: Nat AUC 0.963 (0.903-1.000). Paired DeLong tests of Nat against each published panel on the same samples (Holm-adjusted within cohort and scoring method): GSE144127: vs Berry86 +0.017 (−0.010 to +0.043), p = 0.216, Holm p = 0.647; vs Combined8 −0.003 (−0.015 to +0.009), p = 0.631, Holm p = 1.000; vs Kaforou +0.034 (−0.003 to +0.071), p = 0.074, Holm p = 0.297; vs S11_MI_meanrank +0.030 (+0.001 to +0.060), p = 0.046, Holm p = 0.232; vs Sweeney3 +0.048 (+0.005 to +0.091), p = 0.029, Holm p = 0.173; vs Zak16 −0.003 (−0.020 to +0.013), p = 0.703, Holm p = 1.000. GSE152218: vs Berry86 −0.026 (−0.070 to +0.017), p = 0.230, Holm p = 0.921; vs Combined8 +0.016 (−0.003 to +0.035), p = 0.094, Holm p = 0.565; vs Kaforou +0.000 (−0.021 to +0.021), p = 1.000, Holm p = 1.000; vs S11_MI_meanrank +0.018 (−0.023 to +0.060), p = 0.385, Holm p = 1.000; vs Sweeney3 +0.018 (−0.027 to +0.064), p = 0.425, Holm p = 1.000; vs Zak16 +0.037 (−0.007 to +0.081), p = 0.097, Holm p = 0.565. No difference is significant after adjustment, so comparability rather than superiority is claimed; against Zak16, a 16-gene signature, the difference is −0.003 (95% CI −0.020 to +0.013) using five genes. In GSE152218 random three-gene panels reach a median AUC of 0.83 and every panel scores above 0.92, so that cohort shows replication rather than specificity.

Published gene signatures also generalised excellently. Following this order by the panels best performing models scores, Berry86 1.000 (MLP) > Sweeney3 0.976 (singscore) > Kaforou 0.966 (singscore) > Zak16 0.950 (LR)

However, this result has to take into account the task simplicity as well, size-matched random panels performed comparably in many cases, reaching median AUCs between 0.806 and 0.956 depending on panel size, indicating that active TB is associated with widespread transcriptional changes detectable by diverse gene sets even when the selection is random.

Median performance did vary across five scoring approaches, although the divergences between the methods are not very high which is very likely due to the simplicity of the task itself which must be considered while making any statements on performance.

#### ATB vs OD external testing

This task demonstrated decent performance in external tests. Held-out AUC while not comparable to ATB vs LTB was still promising across models and panels with GSE144127 Nat Singscore at (0.861) being the best native result. The other four scoring approaches also performed very well with held-out AUCs being singscore (0.855), MLP (0.853), logistic regression (0.848) and random forest (0.734) (Fig. 7A).

Published gene signatures also generalised to some extent even if at not the level of ATB vs LTB. Following this order by the panels best performing models scores, Zak16 0.865 (MLP) > Sweeney3 0.853 (singscore) > Berry86 0.839 (DirAvg) > Kaforou 0.821 (DirAvg)

Size-matched random panels performed much worse in ATB vs OD task comparatively, reaching median AUCs between 0.498 and 0.559 depending on panel size, indicating that the selected panels do look for a hard to catch but real signal separating the two classes considering the randomly picked panels are not even close performance wise.

Median performance did vary between models much like in the internal cross-cohort analysis adding more evidence that model choice matters specifically for ATB vs OD task, since even within the same panel model choice can cause the AUCs to fluctuate as much as 0.861-0.734, as observed in Nat Singscore vs Nat RF.

#### LTB vs CON external testing

This test was not conducted due to an absence of external test datasets, and low confidence in its performance after its poor performance in internal cross-cohort testing.

### P3: Domain Shift and PCA Analysis results

The structural differences observed between the training and test cohorts are consistent with the concept of dataset shift in machine learning biomarker development (34), in which distributional mismatches between training and target populations degrade classifier performance even when internal validation is strong. PCA revealed task-dependent patterns of cohort structure (**Fig. 8**).

**Fig 8.**
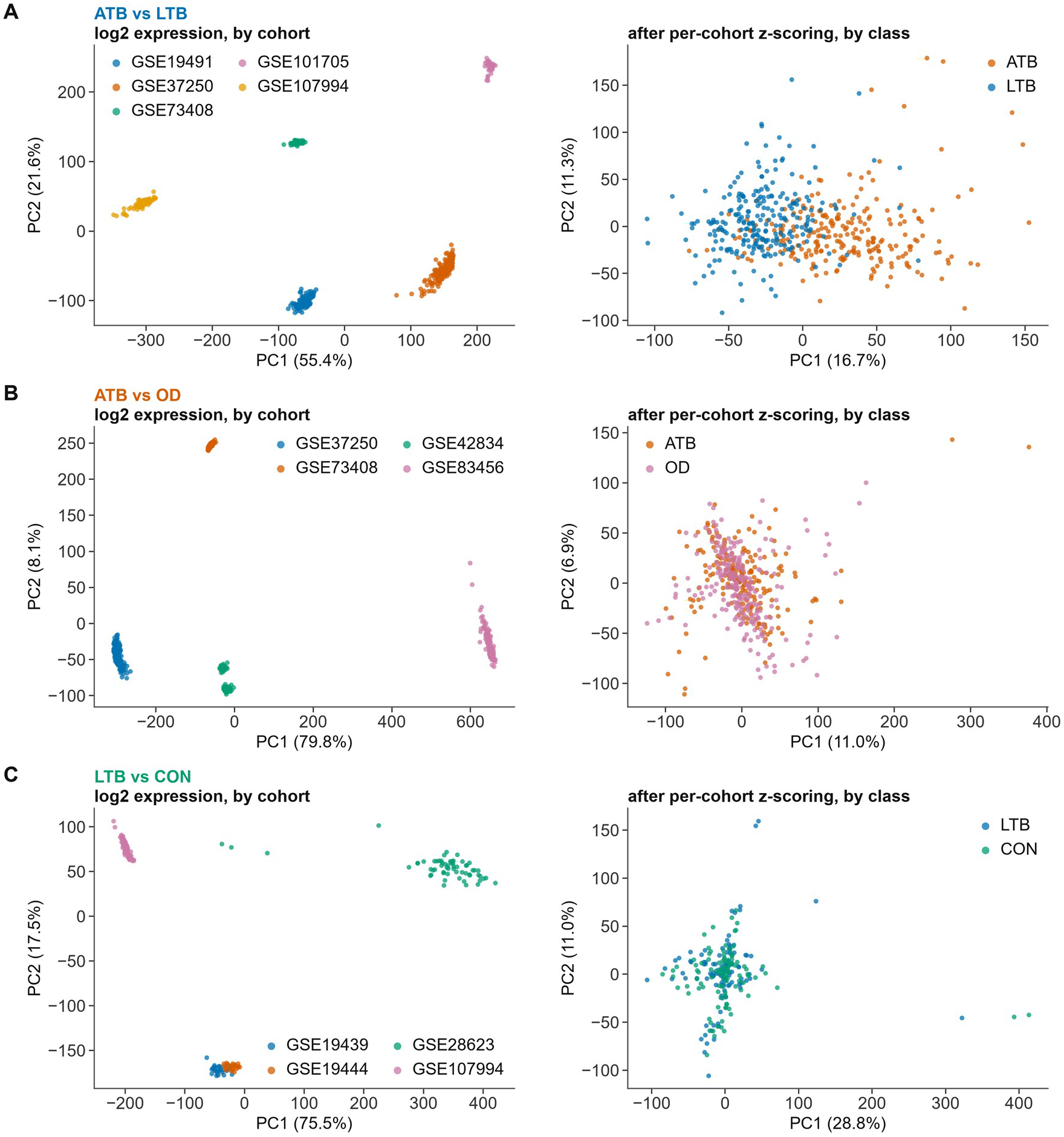
Cohort structure before and after per-cohort normalisation. Principal component analysis of the samples entering each task, computed on that task’s gene universe. Left column: log2 expression before normalisation, coloured by cohort. Right column: the same samples after the label-free per-cohort z-scoring used by every model, coloured by class. Axis labels give the percentage of total variance carried by each component. (A) ATB vs LTB, (B) ATB vs OD, (C) LTB vs CON. Share of the variance in the first two components attributable to cohort and to class (one-way R2): ATB vs LTB: before, cohort R2 = 0.99 and class R2 = 0.01; after, cohort R2 = 0.03 and class R2 = 0.26 ATB vs OD: before, cohort R2 = 1.00 and class R2 = 0.00; after, cohort R2 = 0.01 and class R2 = 0.00 LTB vs CON: before, cohort R2 = 0.97 and class R2 = 0.01; after, cohort R2 = 0.00 and class R2 = 0.00 Cohort identity accounts for almost all of the leading variance before normalisation and almost none after it. Class is not a leading axis of variance in any task, weakly so in ATB vs LTB; in ATB vs OD the class difference is absent from the first two components even though the five-gene signature reaches an AUC of 0.855 in an independent cohort, so unsupervised structure does not track discriminability.

For ATB vs LTB, active TB samples and latent TB formed clusters next to each other, with some overlap but partially distinct clusters. Class explains 25.9% of PC1–PC2 variance after z-scoring (R2 = 0.259), an axis exists but is clearly secondary.

For ATB vs OD, active TB and Other diseases also clustered together but much tighter than the ATB and LTB clouds from the earlier task, there is comparatively more overlap between the two groups too compared to the earlier task. Class explains 0.2% of PC1-PC2 variance after z-scoring (R2 = 0.002). There is no geometric separation.

For LTB vs CON, extensive overlap between classes was observed with distinguishing the two nearly impossible geometrically, as the overlap was worse than even the ATB vs OD task.

As for Domain shift, in the pre normalisation it represents the bulk of the drift as expected, where every dataset clumps separate from each other, with the exception of GSE 19439 and GSE 19444 which as sister sub series clump together, leading to the scepticism towards positive results on either when the training set held the other (**Fig. S8**).

Post normalisation this cohort gap is blunted as shown in the table below for reference.

| Task | Cohort (log2) | Class (log2) | Cohort (z) | Class (z) |
|---|---|---|---|---|
| ATB vs LTB | 0.995 | 0.007 | 0.034 | 0.259 |
| ATB vs OD | 0.998 | 0.001 | 0.015 | 0.002 |
| LTB vs CON | 0.966 | 0.007 | 0.004 | 0.004 |

## Discussion

### Our contribution in this study

Our findings add to the recent work highlighting generalisation challenges in TB transcriptomics (8,9,12). Substantial performance heterogeneity across 16 published signatures was reported, with external validation showing a marked decline in performance (8). Our systematic model comparison extends these observations by demonstrating that model architecture alone cannot overcome cohort-specific structure.

The studies evaluated published signatures as fixed objects. Comparatively speaking in our study every panel was rebuilt inside each fold and trained and tested with LOCO methodology and then post construction from discovered panels scored once on a cohort it had never seen, while comparing it with randomly picked genes functioning as control with size-matched random panels as the reference.

This design allows for selector, model and cohort composition to be separated rather than confounded in each test and has been used to rule out confounds from disease composition, panel origin, model, scoring system and even panel size, with the analysis spanning 13 folds, 3 tasks, 6 panels, 5 scoring methods and 2 external cohorts.

### Task/model Dependence

Task remained the biggest source of difference in performance when all other parameters were held constant: data, folds, models and the random controls. We were also able to demonstrate that class becomes a visible axis of variance within tests only in ATB vs LTB; however, even in that instance it remains a secondary one (R2 = 0.259). The difference in task-by-task median classification performance post normalisation shows this clearly.

| Task | ATB vs LTB | ATB vs OD | LTB vs CON |
|---|---|---|---|
| Median AUC | 0.95 | 0.75 | 0.57 |

Corroboration from another analysis that did not see the data labels further proves this point, (PCA class R2 on PC1-PC2 after per-cohort z-scoring)

| Task | ATB vs LTB | ATB vs OD | LTB vs CON |
|---|---|---|---|
| Class Difference | 0.259 | 0.002 | 0.004 |

The fact that method spread in performance is far smaller than panel spread also indicates architecture does not matter as much as panel choice.

| Spread type | ATB vs LTB | ATB vs OD | LTB vs CON |
|---|---|---|---|
| Method | 0.006 | 0.048 | 0.050 |
| Panel | 0.037 | 0.279 | 0.088 |

Panel choice thus dominates model choice everywhere. ATB vs OD is the only task where architecture matters, and even there MLP’s lead comes with a +0.137 train-to-held-out drop against +0.010 for singscore, as earlier mentioned, its complexity aids it in buying fit, not transfer. In the hardest task the simplest score wins outright. It being LTB v CON where DirAvg is the best method at 0.613 AUC.

### The mRMR Finding

Our comparisons with mRMR methodologies also showed an unexpected result, mRMR derived panels have been used to great effect in transcriptomics and other computational papers before but in our analysis, Nat(mRMR) proved to not be very satisfactory, It held itself well in ATB vs LTB but as previously mentioned, the task simplicity allows all panels to perform well, It however heavily underperformed on ATB vs OD during the tests we performed, it was beaten by random panels in every cell, the scores observed are as follows,

| Type | Median AUC | Range | Beats Random Score |
|---|---|---|---|
| Nat(mRMR) as originally specified | 0.549 | 0.399-0.683 | 0/20 |
| Relevance and redundancy rescaled | 0.614 | 0.484-0.717 | 1/20 |
| MI top-20 union + forward selection (mRMR dropped, wrapper kept) | 0.713 | 0.524-0.814 | 7/20 |
| MI mean-rank, no mRMR, no wrapper | 0.723 | 0.612-0.824 | 10/20 |
| Direction-consistency + forward selection | 0.771 | 0.632-0.827 | 13/20 |
| Nat (forward selection dropped) | 0.786 | 0.641-0.817 | 13/20 |

From our tests, removing the mRMR redundancy step from the pipeline, with the wrapper unchanged, recovers +0.164 of AUC, with it being the single largest gain in performance. In the next step, rescaling the two mRMR terms recovers an additional +0.065 in performance, as such the scale mismatch between relevance and redundancy explains part of the failure. Even so, the penalty remains harmful even when the terms are comparable (S5 Table).

We then tried replacing mutual information with direction-consistency, which resulted in the gain of an additional +0.063 with the wrapper dropped. The forward-selection wrapper never helps transfer, removing it granted an additional +0.010 under mutual information and +0.015 under direction-consistency, while raising the cross-cohort AUC from 0.79 to 0.89.

We presume that this is due to the way the mRMR system works. When the transferable signal is carried by a tightly co-expressed module, much like the ATB interferon response; the various observed genes in the transcriptome change expression in the same manner. This we presume causes the redundancy term to start filtering them out aggressively.

When this process begins and the redundancy term dominates the criterion the selector systematically starts discarding the co-regulated genes that carry the signal, since they qualify as being “redundant” in this context. This causes them to be replaced with mutually uncorrelated genes. Whose individual relevance is marginal and, often very cohort-specific. As a result the list is filled with genes that while not redundant perform much worse qualitatively on a classification panel.

### The Sarcoidosis Finding and its implications

In our ATB vs OD tests, it has been observed that while the Nat panel and other panels are able to differentiate between TB and other diseases such as pneumonia, LRTI/URTI and cancer at a performance of 0.90 - 0.94 AUC, it struggles to differentiate between active TB and sarcoidosis, scoring a mere 0.577 AUC. This crash in performance is not limited to Nat alone but affects every other panel as well, with performance hovering between 0.549 - 0.597 (**S9 Table**).

This finding points to a shared granulomatous interferon programme between sarcoidosis and ATB in whole blood, which are indistinguishable by the methods tested in this paper. This however does not rule out the possibility of the use of the ATB vs OD classifier for clinical testing due to some of the other observations made by us.

The global incidence of Tuberculosis dwarfs that of sarcoidosis: 390,000 new cases of sarcoidosis a year (35) against 10.7 million new cases of tuberculosis in 2024 reported by WHO (2). In South Asia specifically, sarcoidosis incidence is 6.44 per 100,000 (5.65– 7.26), accounting for 0.096M of the 0.39M global cases.

Taken together with a TB incidence of about 187 per 100,000 (2) in the same region, the ratio of sarcoidosis to tuberculosis patients works out at roughly 1:30, with the tuberculosis count dwarfing the sarcoidosis count.

To get a better idea of the possibility of a triage, we would then test the sensitivity and specificity to see if we could meet the WHO triage criteria due to these reasons, our findings were,

|  | **Sensitivity** | **Specificity** |
|---|---|---|
| **ATB v OD, GSE144127** |  |  |
| All samples | 86.1% (77.6–93.2) | 69.3% (63.5–74.5) |
| Sarcoidosis removed | 86.1% | 75.3% (70.2–80.5) |
| Sarcoidosis only | 86.1% | 21.2% (7.1–35.3) |

Note that a triage test is defined by an operating point, so a decision threshold was needed. The threshold was set on the discovery cohorts alone, in this case being pooled LOCO at 90% sensitivity, then applied without change to the validation cohort, so nothing was tuned on validation data.

By comparison ATB v LTB was 92.9% Sensitivity and 74.1% Specificity. Due to this, ATB vs LTB cleared the WHO criteria on point estimate, however ATB vs OD missed both criteria, reaching 86.1% sensitivity against the 90% minimum and 69.3% specificity against 70%. Removing sarcoidosis raised specificity to 75.3% while leaving sensitivity unchanged, so sarcoidosis costs roughly six percentage points of specificity and it is sensitivity that keeps the panel below the triage standard.

Due to these facts despite the shortcomings of the panel, due to the effective sizes of the two groups of patients, and the fact that the error is false positives on sarcoidosis being referred as tuberculosis, instead of vice versa, only a small segment of the population will be getting an incorrect diagnosis, and following this diagnosis a confirmatory test would clear the ailment. On the flipside, the panel will allow for a triage of Tuberculosis from most other ailments which number far greater than the sarcoidosis samples.

## Conclusions

We systematically benchmarked machine learning classifiers for TB transcriptomic classification and observed strongly task-dependent cross-cohort generalisability.

Active TB vs latent TB classification achieved excellent external performance, demonstrating that distinguishing active from latent infection is feasible across diverse populations, likely due to the extremely large shift in the transcriptome across many genes, to the extent that even randomly picked panels performed very well at class discrimination.

In contrast, active TB vs other diseases proved markedly harder within the cross-cohort benchmark, likely driven by cohort-specific disease composition, although the locked panel still transferred to the external cohort at an AUC of 0.855, statistically indistinguishable from every published signature tested. Depending on the other diseases class the performance was either satisfactory or disappointing as in the case of sarcoidosis as mentioned earlier in the text (S8 Table).

Latent TB vs Control meanwhile was a failure with discrimination being completely unsuccessful.

We assessed the possibility of using the classifier for public triage with the findings. With the threshold fixed on discovery data, ATB vs LTB met the WHO triage minimum while ATB vs OD fell short on sensitivity, so the approach is close but not yet at a deployable standard for public use.

We also assessed the performance of mRMR in panel design and its weaknesses in biological contexts where co-expressed redundant genes are the primary discriminating force between classes.

Across tasks, model architecture exerted limited influence on generalisability, with only isolated cohort-specific advantages with the exception of ATB v OD. Differences between panels had comparatively more of an impact on results.

## Limitations

Our analysis was limited to publicly available datasets, which may not represent all TB populations or clinical settings. The exclusion of pediatric cohorts and HIV-coinfected individuals limits generalisability to these important subgroups.

Despite our extensive LOCO and cross-cohort testing true external testing was only done with one dataset each for ATB vs LTB and ATB vs OD and no suitable external cohort existed for the third task. This is a valid criticism of the study design caused due to our willingness to compare panels against more datasets in LOCO and derive a better Nat panel as well as the scarcity of large enough datasets of the kind needed for the study.

The ATB vs LTB validation cohort shares the same geographical region of recruitment with one of the discovery cohorts, and the two accessions cite the same downstream literature. We tested for sample overlap, but none was found, and sample-level expression comparison placed every pair of compared samples variation below the corresponding within-cohort similarity, with cross-cohort resemblance tracking sequencing platform rather than study origin.

Independence of recruitment nonetheless cannot be established from public metadata, combined with the smaller size of the dataset itself totalling at 14 active and 27 latent cases, with a singular female active case its confidence intervals are wide and so this fact must be kept in consideration while reading the results, its also why sex-stratified estimates are not possible.

Additionally, as flagged in the methods section, two cohorts in the LTB vs CON task, GSE19439 and GSE19444, are sub-series of the same superseries, In the LOCO approach whenever one of these was in the testing cohort the other was in train and as observed in the domain shift analysis section and the plot, the two datasets are extremely similar in background and any test results on one when training encompassed the other will give optimistic results. As was predicted the few cells in this task that exceeded their random comparators fall almost entirely in these two folds.

Additionally, multiplicity was addressed only in the external validation cohorts, where panels were compared by Holm-adjusted paired DeLong tests (1). Within the rotating benchmark however, panels were compared only against their own size-matched random distributions and no adjusted p-values are reported. This comparison for selectors, however, was done post-hoc after observation of mRMR’s failure to classify. That being said, no test data leaked into selection as each variant was rebuilt inside every fold and only scored on the held-out cohorts.

Since the final panels were also derived from the cohorts that underwent LOCO, the final Nat panels, of three and five genes, did not undergo any testing other than the external tests, this can be observed in the supplementary table.

The sarcoidosis comparison performed by us also rests on 79 active TB against 33 sarcoidosis cases in a single cohort. The result was an AUC of 0.577 with a 95% interval of 0.46–0.70 that includes chance. As such the size of the effect is not well estimated, more datasets comprising for TB and Sarcoidosis samples will be needed to acquire more accurate figures.

In our study we also presume reported sarcoidosis incidence is indicative of true incidence. However, the possibility exists that in high-TB-burden settings; sarcoidosis is plausibly under-diagnosed due to lower penetration of health services, or misdiagnosed as tuberculosis itself, which is the same confusion the panels exhibit.

Our use of binary classification tasks (ATB vs LTB, etc.) does not capture the continuous spectrum of TB infection states. More nuanced phenotypic definitions may be needed for clinically useful signatures that target the progression from latent to active TB.

We evaluated fixed gene panels rather than exploring adaptive or transfer learning approaches that might improve cross-cohort generalisation. Future work should investigate whether cohort-aware modelling strategies can mitigate the domain shift we observed as it can vastly improve model performances for diagnostics.

## Supporting information

S1 Fig. ATB vs LTB: every panel against size-matched random panels

S1 Table. ATB vs LTB: cross-cohort benchmark, all panels and scoring methods

S2 Fig. ATB vs OD: every panel against size-matched random panels

S2 Table. ATB vs OD: cross-cohort benchmark, all panels and scoring methods

S3 Fig. LTB vs CON: every panel against size-matched random panels

S3 Table. LTB vs CON: cross-cohort benchmark, all panels and scoring methods

S4 Fig. ATB vs LTB: derivation of Nat under all five scoring methods

S4 Table. Nat panel size selected in each fold by the inner leave-one-cohort-out loop

S5 Fig. ATB vs OD: derivation of Nat under all five scoring methods

S5 Table. Comparison of six feature-selection criteria

S6 Fig. LTB vs CON: derivation of Nat under all five scoring methods

S6 Table. Frozen Nat panels and the recurrence of their genes across folds

S7 Fig. Every gene selected by the mRMR-derived panel, by fold

S7 Table. External validation: locked panels applied without refitting

S8 Fig. Cohort-level offset diagnostic

S8 Table. Paired DeLong comparisons in the external cohorts

S9 Table. Subgroup analyses in the external cohorts (directional average)

S10 Table. Direction preservation and leakage canary in the external cohorts

## Declarations

### Author Contributions

**BA:** Conceptualization, Methodology, Software, Formal analysis, Investigation, Data curation, Writing - original draft, Visualization. **SKP:** Writing - review and editing, Methodology, Visualization. **BT:** Supervision, Writing - review and editing, Project administration.

## Ethics Approval and Consent to Participate

Ethics approval was not required as all data were obtained from publicly available repositories with no patient identifiers.

## Data Availability

All datasets analysed are publicly available from the Gene Expression Omnibus (GEO): GSE19491, GSE37250, GSE73408, GSE19439, GSE19444, GSE28623, GSE101705, GSE107994, GSE42834, GSE83456, and the validation cohorts GSE144127 and GSE152218. Processed datasets are deposited at Zenodo https://doi.org/10.5281/zenodo.18923347. The deposit contains the source input matrices as well as the processed datasets, so the analysis scripts run end to end from a single download. The analysis scripts used for the same are available at https://github.com/Brendan-Alex/Analysis-scripts-used-for-TB-transcriptomics

## Acknowledgements

We thank Rahul K from IIT Hyderabad for methodological review support during the early stages of this project.

## Declaration of generative AI and AI-assisted technologies in the manuscript preparation process

During the preparation of this work, the authors used GitHub Copilot (Microsoft Corporation) to assist with the scripts that retrieved and assembled the source expression data; Claude (Anthropic PBC), used via its Cowork agentic interface, to write, debug and run the analysis and plotting code, to generate the figures, to draft the Methods section of the paper, to check reported values within the manuscript against the analysis output files, to correct typographical errors and cross-references, and to revise individual sentences for factual accuracy; and Grammarly GO (Grammarly Inc.) for grammar and language correction. The Abstract, Results, Discussion and Conclusions were entirely written by the authors. The authors specified every analysis, inspected the results, and reviewed and edited all tool-generated content, and take full responsibility for the published article. All analysis code is available at the repository listed under Data Availability.

## Supporting information

**S1 Fig. ATB vs LTB: every panel against size-matched random panels, all five scoring methods.** Rows are scoring methods; groups along the x-axis are held-out cohorts. Each point is the held-out AUC with its bootstrap 95% confidence interval (1,000 resamples, seed 42), drawn over the AUCs of the 50 random gene panels of the same size in the same fold (light grey = 2.5th-97.5th percentile, dark grey = interquartile range, white line = median). Filled circles are clean cells; open diamonds mark derivation overlap with the held-out cohort and open squares the GSE19439 sibling fold. An asterisk marks clean cells where at most 5% of random panels reached the panel’s AUC. Nat is the fold-level panel at the size chosen for that fold and method. Cells beating random, all methods: Nat 20/25 clean; Nat(mRMR) 14/25 clean; Zak16 5/15 clean; Kaforou 7/15 clean; Berry86 11/20 clean; Sweeney3 11/15 clean.

**S2 Fig. ATB vs OD: every panel against size-matched random panels, all five scoring methods.** Rows are scoring methods; groups along the x-axis are held-out cohorts. Each point is the held-out AUC with its bootstrap 95% confidence interval (1,000 resamples, seed 42), drawn over the AUCs of the 50 random gene panels of the same size in the same fold (light grey = 2.5th-97.5th percentile, dark grey = interquartile range, white line = median). Filled circles are clean cells; open diamonds mark derivation overlap with the held-out cohort and open squares the GSE19439 sibling fold. An asterisk marks clean cells where at most 5% of random panels reached the panel’s AUC. Nat is the fold-level panel at the size chosen for that fold and method. Cells beating random, all methods: Nat 15/20 clean; Nat(mRMR) 0/20 clean; Zak16 7/10 clean; Kaforou 14/15 clean; Berry86 13/20 clean; Sweeney3 8/10 clean.

**S3 Fig. LTB vs CON: every panel against size-matched random panels, all five scoring methods.** Rows are scoring methods; groups along the x-axis are held-out cohorts. Each point is the held-out AUC with its bootstrap 95% confidence interval (1,000 resamples, seed 42), drawn over the AUCs of the 50 random gene panels of the same size in the same fold (light grey = 2.5th-97.5th percentile, dark grey = interquartile range, white line = median). Filled circles are clean cells; open diamonds mark derivation overlap with the held-out cohort and open squares the GSE19439 sibling fold. An asterisk marks clean cells where at most 5% of random panels reached the panel’s AUC. Nat is the fold-level panel at the size chosen for that fold and method. Cells beating random, all methods: Nat 4/15 clean; Nat(mRMR) 0/15 clean; Zak16 0/10 clean; Berry86 0/10 clean; Sweeney3 0/5 clean.

**S4 Fig. ATB vs LTB: derivation of Nat under all five scoring methods.** (A) Rows are held-out cohorts and columns scoring methods. The coloured line is the held-out AUC of the top k genes of that fold’s training-only consistency ranking, over the AUCs of 50 random panels of the same size in the same fold (shading as in S1-S3 Figs). The open circle and dotted line mark the size chosen by the inner leave-one-cohort-out loop, which is chosen separately for each method. (B) Final all-cohort derivation: mean inner leave-one-cohort-out AUC against the gene added at each step of the nested ranking. This is an inner-loop estimate over training cohorts, not held-out performance, and is not comparable with panel A. Chosen k per fold: Logistic regression: GSE37250 3, GSE19491 3, GSE107994 2, GSE73408 1, GSE101705 8 | Random forest: GSE37250 8, GSE19491 9, GSE107994 13, GSE73408 13, GSE101705 7 | MLP: GSE37250 6, GSE19491 3, GSE107994 3, GSE73408 1, GSE101705 4 | singscore: GSE37250 13, GSE19491 3, GSE107994 2, GSE73408 1, GSE101705 8 | DirAvg: GSE37250 6, GSE19491 8, GSE107994 3, GSE73408 1, GSE101705 4.

**S5 Fig. ATB vs OD: derivation of Nat under all five scoring methods.** (A) Rows are held-out cohorts and columns scoring methods. The coloured line is the held-out AUC of the top k genes of that fold’s training-only consistency ranking, over the AUCs of 50 random panels of the same size in the same fold (shading as in S1-S3 Figs). The open circle and dotted line mark the size chosen by the inner leave-one-cohort-out loop, which is chosen separately for each method. (B) Final all-cohort derivation: mean inner leave-one-cohort-out AUC against the gene added at each step of the nested ranking. This is an inner-loop estimate over training cohorts, not held-out performance, and is not comparable with panel A. In ATB vs OD the untrained scores need few genes (final k = 5 for both the directional average and singscore) while the trained models prefer long panels (final k: logistic regression 17, random forest 19, MLP 18), so the plateau depends on the scoring method. Chosen k per fold: Logistic regression: GSE37250 11, GSE73408 1, GSE42834 7, GSE83456 9 | Random forest: GSE37250 17, GSE73408 17, GSE42834 7, GSE83456 13 | MLP: GSE37250 17, GSE73408 17, GSE42834 2, GSE83456 9 | singscore: GSE37250 16, GSE73408 11, GSE42834 18, GSE83456 9 | DirAvg: GSE37250 16, GSE73408 12, GSE42834 10, GSE83456 5.

**S6 Fig. LTB vs CON: derivation of Nat under all five scoring methods.** (A) Rows are held-out cohorts and columns scoring methods. The coloured line is the held-out AUC of the top k genes of that fold’s training-only consistency ranking, over the AUCs of 50 random panels of the same size in the same fold (shading as in S1-S3 Figs). The open circle and dotted line mark the size chosen by the inner leave-one-cohort-out loop, which is chosen separately for each method. (B) Final all-cohort derivation: mean inner leave-one-cohort-out AUC against the gene added at each step of the nested ranking. This is an inner-loop estimate over training cohorts, not held-out performance, and is not comparable with panel A. Chosen k per fold: Logistic regression: GSE19439 6, GSE19444 6, GSE28623 3, GSE107994 3 | Random forest: GSE19439 6, GSE19444 2, GSE28623 20, GSE107994 9 | MLP: GSE19439 6, GSE19444 6, GSE28623 3, GSE107994 3 | singscore: GSE19439 1, GSE19444 6, GSE28623 3, GSE107994 3 | DirAvg: GSE19439 10, GSE19444 20, GSE28623 19, GSE107994 3.

**S7 Fig. Every gene selected by the mRMR-derived panel, by fold.** Presence (blue) of each gene in each fold’s 20-gene Nat(mRMR) panel; columns are held-out cohorts and the number at right is the count of folds selecting that gene. Sex-chromosome genes are excluded from selection. Coloured names are HGNC-flagged entries. Summary: ATB vs LTB 53 distinct genes, 0 in every fold, pairwise fold overlap 2– 11 genes; ATB vs OD 55 distinct genes, 0 in every fold, pairwise fold overlap 3–10 genes; LTB vs CON 46 distinct genes, 0 in every fold, pairwise fold overlap 4–8 genes.

**S8 Fig. Cohort-level offset diagnostic.** AUC of the per-sample mean across all measured genes for the positive class, with a bootstrap 95% confidence interval (1,000 resamples). Values at right give the AUC and, in brackets, the share of genes individually separating the classes at |AUC - 0.5| > 0.2. A value of 0.5 indicates no cohort-level offset aligned with class. Open diamonds mark quantile-normalised cohorts (GSE19491 and GSE42834 as deposited, GSE83456 rebuilt in Stage7 with log2 and quantile normalisation), where this diagnostic is near-constant by construction. Range 0.193 to 0.657. Source: Stage7/results/global_shift_heldout.csv.

**S1 Table. ATB vs LTB: cross-cohort benchmark, all panels and scoring methods.** AUC in the held-out cohort for every panel and scoring method. Nat was rebuilt in each fold and its size chosen by an inner leave-one-cohort-out loop separately for each method, so the gene count is given as a range across the five methods; all other panels are fixed. The 95% confidence interval is given for the directional average, the primary score; intervals for the other methods are in the deposited analysis outputs. Random median is the median AUC of 50 size-matched random panels drawn for that fold and panel size. Beats random counts the scoring methods whose AUC was reached by at most 5% of those random panels. “Derivation overlap” marks cells where the published panel’s own derivation cohort, or one of its sub-series, is the held-out cohort; those AUCs are optimistic. Sibling sub-series marks the GSE19439 fold, whose training cohorts include its sister sub-series GSE19444. Values are not pooled or averaged across folds. Nat (per-fold) is not the same object as the frozen Nat (final) panel evaluated in S7-S10 Tables, which was derived once on all cohorts and therefore cannot be scored on any discovery cohort.

**S2 Table. ATB vs OD: cross-cohort benchmark, all panels and scoring methods.** AUC in the held-out cohort for every panel and scoring method. Nat was rebuilt in each fold and its size chosen by an inner leave-one-cohort-out loop separately for each method, so the gene count is given as a range across the five methods; all other panels are fixed. The 95% confidence interval is given for the directional average, the primary score; intervals for the other methods are in the deposited analysis outputs. Random median is the median AUC of 50 size-matched random panels drawn for that fold and panel size. Beats random counts the scoring methods whose AUC was reached by at most 5% of those random panels. Derivation overlap marks cells where the published panel’s own derivation cohort, or one of its sub-series, is the held-out cohort; those AUCs are optimistic. Sibling sub-series marks the GSE19439 fold, whose training cohorts include its sister sub-series GSE19444. Values are not pooled or averaged across folds. Nat (per-fold) is not the same object as the frozen Nat (final) panel evaluated in S7-S10 Tables, which was derived once on all cohorts and therefore cannot be scored on any discovery cohort.

**S3 Table. LTB vs CON: cross-cohort benchmark, all panels and scoring methods.**AUC in the held-out cohort for every panel and scoring method. Nat was rebuilt in each fold and its size chosen by an inner leave-one-cohort-out loop separately for each method, so the gene count is given as a range across the five methods; all other panels are fixed. The 95% confidence interval is given for the directional average, the primary score; intervals for the other methods are in the deposited analysis outputs. Random median is the median AUC of 50 size-matched random panels drawn for that fold and panel size. Beats random counts the scoring methods whose AUC was reached by at most 5% of those random panels. Derivation overlap marks cells where the published panel’s own derivation cohort, or one of its sub-series, is the held-out cohort; those AUCs are optimistic. Sibling sub-series marks the GSE19439 fold, whose training cohorts include its sister sub-series GSE19444. Values are not pooled or averaged across folds. Nat (per-fold) is not the same object as the frozen Nat (final) panel evaluated in S7-S10 Tables, which was derived once on all cohorts and therefore cannot be scored on any discovery cohort.

**S4 Table. Nat panel size selected in each fold by the inner leave-one-cohort-out loop.** For each fold the direction-consistency ranking was rebuilt on the training cohorts only, and k (1–20) was chosen as the smallest value whose mean inner-loop AUC was within 0.01 of the best, separately for each scoring method. Within a fold the ranking is identical for all scoring methods, which differ only in where it is cut; the panel selected in each fold is the first k genes of that fold’s ranking, and the full per-fold rankings are in the deposited analysis outputs. The final column is the held-out AUC at the chosen k minus the held-out AUC a fixed 20-gene panel would have given in the same fold.

**S5 Table. Comparison of six feature-selection criteria.** Every variant builds a 20-gene panel per fold, is rebuilt inside the fold using the training cohorts only, and is scored once on the held-out cohort. Median, minimum and maximum are taken over all fold-by-method cells of the task (25 for ATB vs LTB, 20 for the other two). Beats random counts the cells whose AUC was reached by at most 5% of 50 size-matched random panels. Note that these are fixed 20-gene panels, so the Nat row here is not identical to the inner-loop-sized Nat reported in S1-S4 Tables.

**S6 Table. Frozen Nat panels and the recurrence of their genes across folds.**The direction-consistency procedure was run once on all discovery cohorts of each task to produce a single frozen panel for deposit and external validation. These panels were derived using every discovery cohort and so cannot be evaluated by leave-one-cohort-out; their only out-of-sample evidence is the external cohorts in S7-S10 Tables. As an internal consistency check, the two right-hand columns count how often each frozen gene was independently recovered by the per-fold derivations reported in S1-S4 Tables: whether it entered that fold’s top-20 direction-consistent ranking, and whether it entered that fold’s selected panel under the directional average. Combined8, used in S7-S9 Tables, is the union of the ATB vs LTB and ATB vs OD panels and contains all eight genes listed here for those two tasks. No panel is proposed for LTB vs CON: the procedure returns eleven genes, but four of them were recovered by no fold at all.

**S7 Table. External validation: locked panels applied without refitting.**GSE144127 (ATB vs OD; 79 active TB vs 300 other disease) and GSE152218 (ATB vs LTB; 14 vs 27). Neither cohort took part in panel derivation, model training or any earlier analysis. Models were trained on all discovery cohorts of the task and scored once. Nat (final) is the frozen panel for that task, derived once on all discovery cohorts of the task and applied here without modification: ANKRD22, FCGR1A and PSTPIP2 for ATB vs LTB (k = 3) and APOL2, WDFY1, BATF2, POLB and GBP2 for ATB vs OD (k = 5). Combined8 is the union of those two panels, an eight-gene set applied unchanged to both tasks. MI mean rank is the corresponding frozen panel from the MI mean-rank selector variant. Random median and the empirical p-value compare the panel with 50 size-matched random panels drawn from the same gene universe. These were drawn only for the panels derived in this study, since the question they address is whether our own selection beats chance; the published panels are compared with one another in S8 Table instead. Empirical p is (number of random panels reaching the observed AUC + 1) / 51, so 0.020 is the smallest value obtainable with 50 draws and is a floor rather than a point estimate. These p-values are not adjusted for multiplicity and should be read across the five scoring methods of a panel together, not singly. The first half of the table represents ATB v OD external testing (GSE 144127), the second half represents ATB v LTB external testing (GSE 152218)

**S8 Table. Paired DeLong comparisons in the external cohorts.** Nat (final) against every other panel, computed on identical samples. ΔAUC is Nat (final) minus comparator. p-values are adjusted by the Holm procedure within each cohort and scoring method, over the six comparators listed. No comparison reaches significance after adjustment in either cohort.

**S9 Table. Subgroup analyses in the external cohorts (directional average).** AUC of each panel within clinical and demographic subgroups of the validation cohorts. The active TB group is held constant and the comparison group is restricted. Sample sizes are given as active TB versus comparator. The GSE152218 female subgroup contains a single active TB case and no AUC is estimable.

**S10 Table. Direction preservation and leakage canary in the external cohorts.** Upper block: for every gene of the locked panels, the direction of change learned in the discovery cohorts, the per-cohort discovery AUC, and the AUC of that single gene in the validation cohort. Lower block: AUC when the validation expression matrix is replaced by noise, with labels unchanged.

## References

1. Holm S. A simple sequentially rejective multiple test procedure. Scand J Stat. 1979;6(2):65– 70.

2. World Health Organization. Global tuberculosis report 2025 [Internet]. Geneva: World Health Organization; 2025. Available from: https://www.who.int/teams/global-programme-on-tuberculosis-and-lung-health/tb-reports/global-tuberculosis-report-2025

3. Cohen A, Mathiasen VD, Schön T, Wejse C. The global prevalence of latent tuberculosis: a systematic review and meta-analysis. Eur Respir J. 2019 Sep;54(3):1900655. doi:10.1183/13993003.00655-2019 PubMed PMID: 31221810.

4. World Health Organization. Tuberculosis [Internet]. 2024. Available from: https://www.who.int/news-room/fact-sheets/detail/tuberculosis

5. Berry MPR, Graham CM, McNab FW, Xu Z, Bloch SAA, Oni T, et al. An interferon-inducible neutrophil-driven blood transcriptional signature in human tuberculosis. Nature. 2010 Aug 19;466(7309):973–7. doi:10.1038/nature09247 PubMed PMID: 20725040; PubMed Central PMCID: PMC3492754.

6. Zak DE, Penn-Nicholson A, Scriba TJ, Thompson E, Suliman S, Amon LM, et al. A prospective blood RNA signature for tuberculosis disease risk. Lancet Lond Engl. 2016 Jun 4;387(10035):2312–22. doi:10.1016/S0140-6736(15)01316-1 PubMed PMID: 27017310; PubMed Central PMCID: PMC5392204.

7. Sweeney TE, Braviak L, Tato CM, Khatri P. Genome-wide expression for diagnosis of pulmonary tuberculosis: a multicohort analysis. Lancet Respir Med. 2016 Mar;4(3):213–24. doi:10.1016/S2213-2600(16)00048-5 PubMed PMID: 26907218; PubMed Central PMCID: PMC4838193.

8. Warsinske H, Vashisht R, Khatri P. Host-response-based gene signatures for tuberculosis diagnosis: A systematic comparison of 16 signatures. PLoS Med. 2019 Apr;16(4):e1002786. doi:10.1371/journal.pmed.1002786 PubMed PMID: 31013272; PubMed Central PMCID: PMC6478271.

9. Vargas R, Abbott L, Bower D, Frahm N, Shaffer M, Yu WH. Gene signature discovery and systematic validation across diverse clinical cohorts for TB prognosis and response to treatment. PLoS Comput Biol. 2023 Jul;19(7):e1010770. doi:10.1371/journal.pcbi.1010770 PubMed PMID: 37471455; PubMed Central PMCID: PMC10393163.

10. Battle A, Mostafavi S, Zhu X, Potash JB, Weissman MM, McCormick C, et al. Characterizing the genetic basis of transcriptome diversity through RNA-sequencing of 922 individuals. Genome Res. 2014 Jan;24(1):14–24. doi:10.1101/gr.155192.113 PubMed PMID: 24092820; PubMed Central PMCID: PMC3875855.

11. Lee MN, Ye C, Villani AC, Raj T, Li W, Eisenhaure TM, et al. Common genetic variants modulate pathogen-sensing responses in human dendritic cells. Science. 2014 Mar 7;343(6175):1246980. doi:10.1126/science.1246980 PubMed PMID: 24604203; PubMed Central PMCID: PMC4124741.

12. Wang X, Harper K, Sinha P, Johnson WE, Patil P. Analysis of the cross-study replicability of tuberculosis gene signatures using 49 curated human transcriptomic datasets. Tuberculosis. 2025 Jul;153:102649. doi:10.1016/j.tube.2025.102649 PubMed PMID: 40359654; PubMed Central PMCID: PMC12158633.

13. Bobak CA, Titus AJ, Hill JE. Comparison of common machine learning models for classification of tuberculosis using transcriptional biomarkers from integrated datasets. Appl Soft Comput. 2019 Jan;74:264–73. doi:10.1016/j.asoc.2018.10.005

14. Kaforou M, Wright VJ, Oni T, French N, Anderson ST, Bangani N, et al. Detection of tuberculosis in HIV-infected and -uninfected African adults using whole blood RNA expression signatures: a case-control study. PLoS Med. 2013 Oct;10(10):e1001538. doi:10.1371/journal.pmed.1001538 PubMed PMID: 24167453; PubMed Central PMCID: PMC3805485.

15. Walter ND, Miller MA, Vasquez J, Weiner M, Chapman A, Engle M, et al. Blood Transcriptional Biomarkers for Active Tuberculosis among Patients in the United States: a Case-Control Study with Systematic Cross-Classifier Evaluation. J Clin Microbiol. 2016 Feb;54(2):274–82. doi:10.1128/JCM.01990-15 PubMed PMID: 26582831; PubMed Central PMCID: PMC4733166.

16. Maertzdorf J, Repsilber D, Parida SK, Stanley K, Roberts T, Black G, et al. Human gene expression profiles of susceptibility and resistance in tuberculosis. Genes Immun. 2011 Jan;12(1):15–22. doi:10.1038/gene.2010.51 PubMed PMID: 20861863.

17. Leong S, Zhao Y, Joseph NM, Hochberg NS, Sarkar S, Pleskunas J, et al. Existing blood transcriptional classifiers accurately discriminate active tuberculosis from latent infection in individuals from south India. Tuberculosis. 2018 Mar;109:41–51. doi:10.1016/j.tube.2018.01.002

18. Singhania A, Verma R, Graham CM, Lee J, Tran T, Richardson M, et al. A modular transcriptional signature identifies phenotypic heterogeneity of human tuberculosis infection. Nat Commun. 2018 Jun 19;9(1):2308. doi:10.1038/s41467-018-04579-w PubMed PMID: 29921861; PubMed Central PMCID: PMC6008327.

19. Bloom CI, Graham CM, Berry MPR, Rozakeas F, Redford PS, Wang Y, et al. Transcriptional Blood Signatures Distinguish Pulmonary Tuberculosis, Pulmonary Sarcoidosis, Pneumonias and Lung Cancers. Rengarajan J, editor. PLoS ONE. 2013 Aug 5;8(8):e70630. doi:10.1371/journal.pone.0070630

20. Blankley S, Graham CM, Turner J, Berry MPR, Bloom CI, Xu Z, et al. The Transcriptional Signature of Active Tuberculosis Reflects Symptom Status in Extra-Pulmonary and Pulmonary Tuberculosis. PloS One. 2016;11(10):e0162220. doi:10.1371/journal.pone.0162220 PubMed PMID: 27706152; PubMed Central PMCID: PMC5051928.

21. Anderson ST, Kaforou M, Brent AJ, Wright VJ, Banwell CM, Chagaluka G, et al. Diagnosis of childhood tuberculosis and host RNA expression in Africa. N Engl J Med. 2014 May 1;370(18):1712–23. doi:10.1056/NEJMoa1303657 PubMed PMID: 24785206; PubMed Central PMCID: PMC4069985.

22. Love MI, Huber W, Anders S. Moderated estimation of fold change and dispersion for RNA-seq data with DESeq2. Genome Biol. 2014 Dec 5;15(12):550. doi:10.1186/s13059-014-0550-8

23. Murchan P, Ó Broin, Baird AM, Sheils O, Finn S. Deep feature batch correction using ComBat for machine learning applications in computational pathology. J Pathol Inform. 2024 Dec 1;15:100396. doi:10.1016/j.jpi.2024.100396

24. Ding C, Peng H. Minimum redundancy feature selection from microarray gene expression data. J Bioinform Comput Biol. 2005 Apr;3(2):185–205. doi:10.1142/s0219720005001004 PubMed PMID: 15852500.

25. Kraskov A, Stögbauer H, Grassberger P. Estimating mutual information. Phys Rev E. 2004 Jun 23;69(6):066138. doi:10.1103/PhysRevE.69.066138

26. Libbrecht MW, Noble WS. Machine learning applications in genetics and genomics. Nat Rev Genet. 2015 Jun;16(6):321–32. doi:10.1038/nrg3920 PubMed PMID: 25948244; PubMed Central PMCID: PMC5204302.

27. Foroutan M, Bhuva DD, Lyu R, Horan K, Cursons J, Davis MJ. Single sample scoring of molecular phenotypes. BMC Bioinformatics. 2018 Dec;19(1):404. doi:10.1186/s12859-018-2435-4

28. Pedregosa F, Varoquaux G, Gramfort A, Michel V, Thirion B, Grisel O, et al. Scikit-learn: Machine Learning in Python. J Mach Learn Res. 2011;12(85):2825–30.

29. Li Y, Yuan Y. Convergence Analysis of Two-layer Neural Networks with ReLU Activation. In: Advances in Neural Information Processing Systems [Internet]. Curran Associates, Inc.; 2017 [cited 2026 Aug 14]. Available from: https://proceedings.neurips.cc/paper_files/paper/2017/hash/a96b65a721e561e1e3de768ac 819ffbb-Abstract.html

30. Suthaharan S. Decision Tree Learning. In: Suthaharan S, editor. Machine Learning Models and Algorithms for Big Data Classification: Thinking with Examples for Effective Learning [Internet]. Boston, MA: Springer US; 2016 [cited 2026 Aug 14]. p. 237–69. Available from: doi:10.1007/978-1-4899-7641-3_10

31. Hoang LT, Jain P, Pillay TD, Tolosa-Wright M, Niazi U, Takwoingi Y, et al. Transcriptomic signatures for diagnosing tuberculosis in clinical practice: a prospective, multicentre cohort study. Lancet Infect Dis. 2021 Mar;21(3):366–75. doi:10.1016/S1473-3099(20)30928-2

32. Johnson WE, Odom A, Cintron C, Muthaiah M, Knudsen S, Joseph N, et al. Comparing tuberculosis gene signatures in malnourished individuals using the TBSignatureProfiler. BMC Infect Dis. 2021 Dec;21(1):106. doi:10.1186/s12879-020-05598-z

33. DeLong ER, DeLong DM, Clarke-Pearson DL. Comparing the Areas under Two or More Correlated Receiver Operating Characteristic Curves: A Nonparametric Approach. Biometrics. 1988 Sep;44(3):837. doi:10.2307/2531595

34. Dockès J, Varoquaux G, Poline JB. Preventing dataset shift from breaking machine-learning biomarkers. GigaScience. 2021 Sep 28;10(9):giab055. doi:10.1093/gigascience/giab055 PubMed PMID: 34585237; PubMed Central PMCID: PMC8478611.

35. Cao J, Li H, Yin X, Yang J, Pu L, Yang J. Global burden of pulmonary sarcoidosis from 1990 to 2021: a comprehensive analysis based on the GBD 2021 study. Front Med. 2025 May 16;12:1585005. doi:10.3389/fmed.2025.1585005

