## Supplementary figures and images for "Cross-cohort benchmarking and validation reveals task-dependent generalisability in tuberculosis blood transcriptomic classification"

### S1 Fig. ATB vs LTB: every panel against size-matched random panels

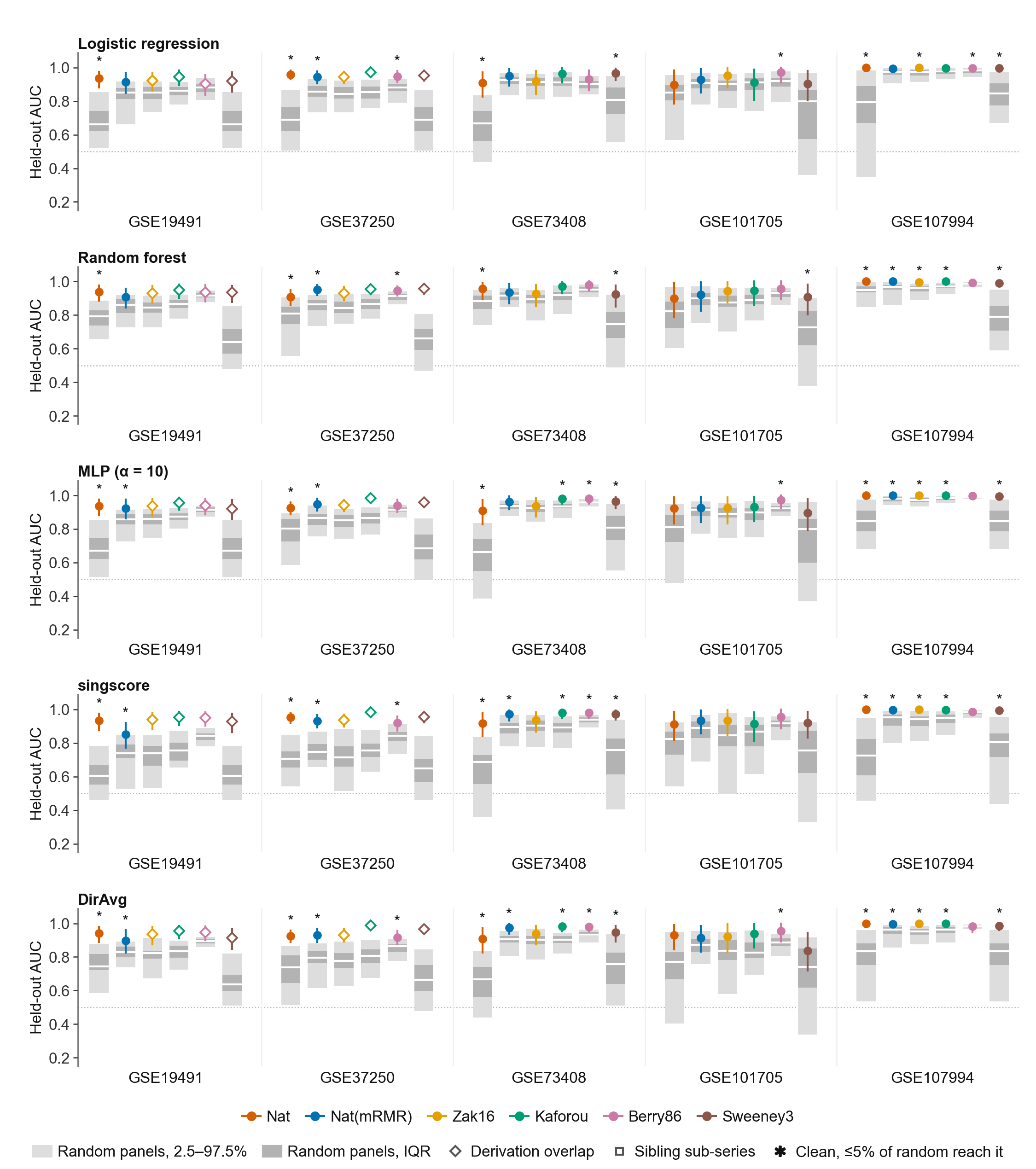

### S2 Fig. ATB vs OD: every panel against size-matched random panels

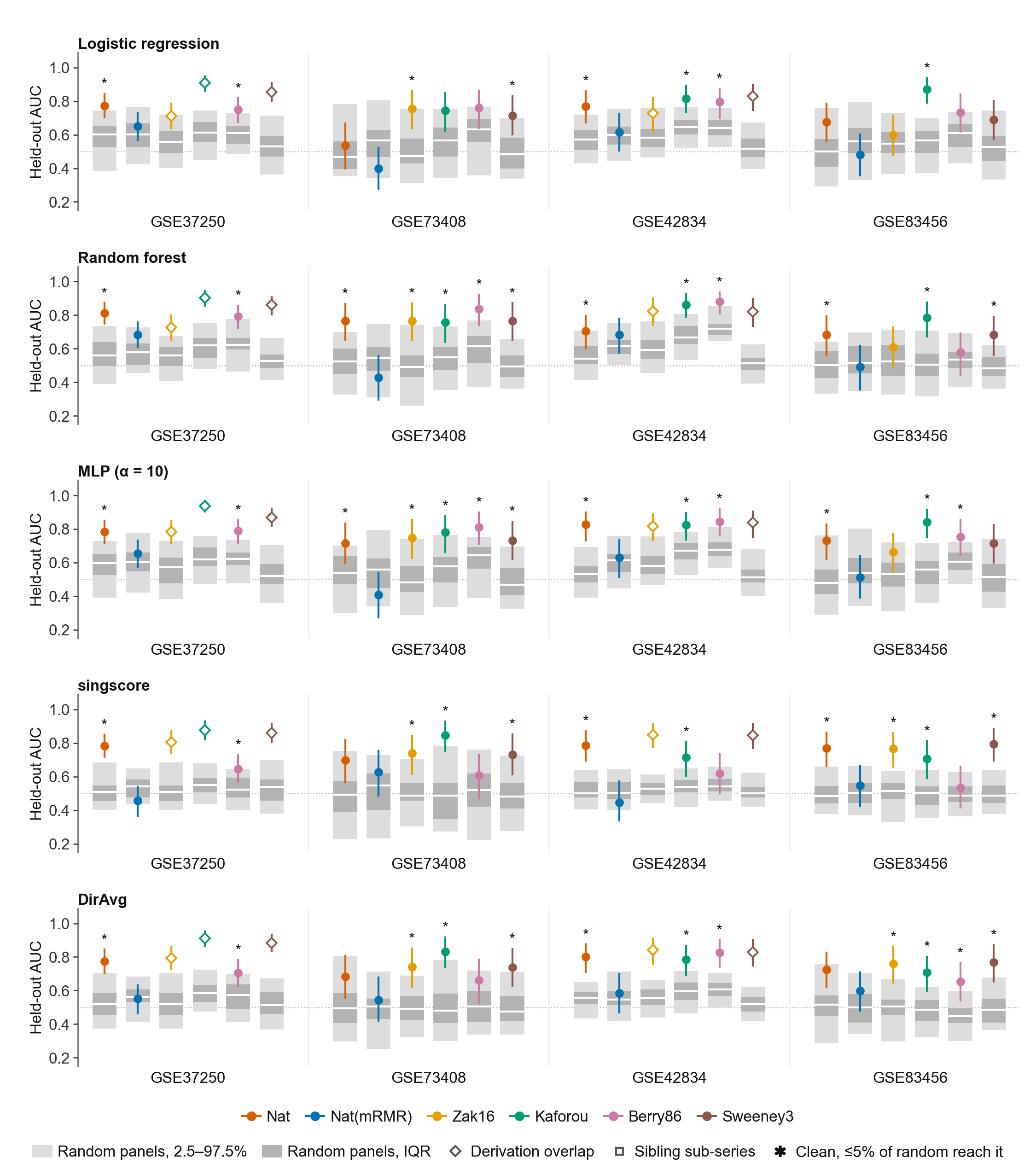

### S3 Fig. LTB vs CON: every panel against size-matched random panels

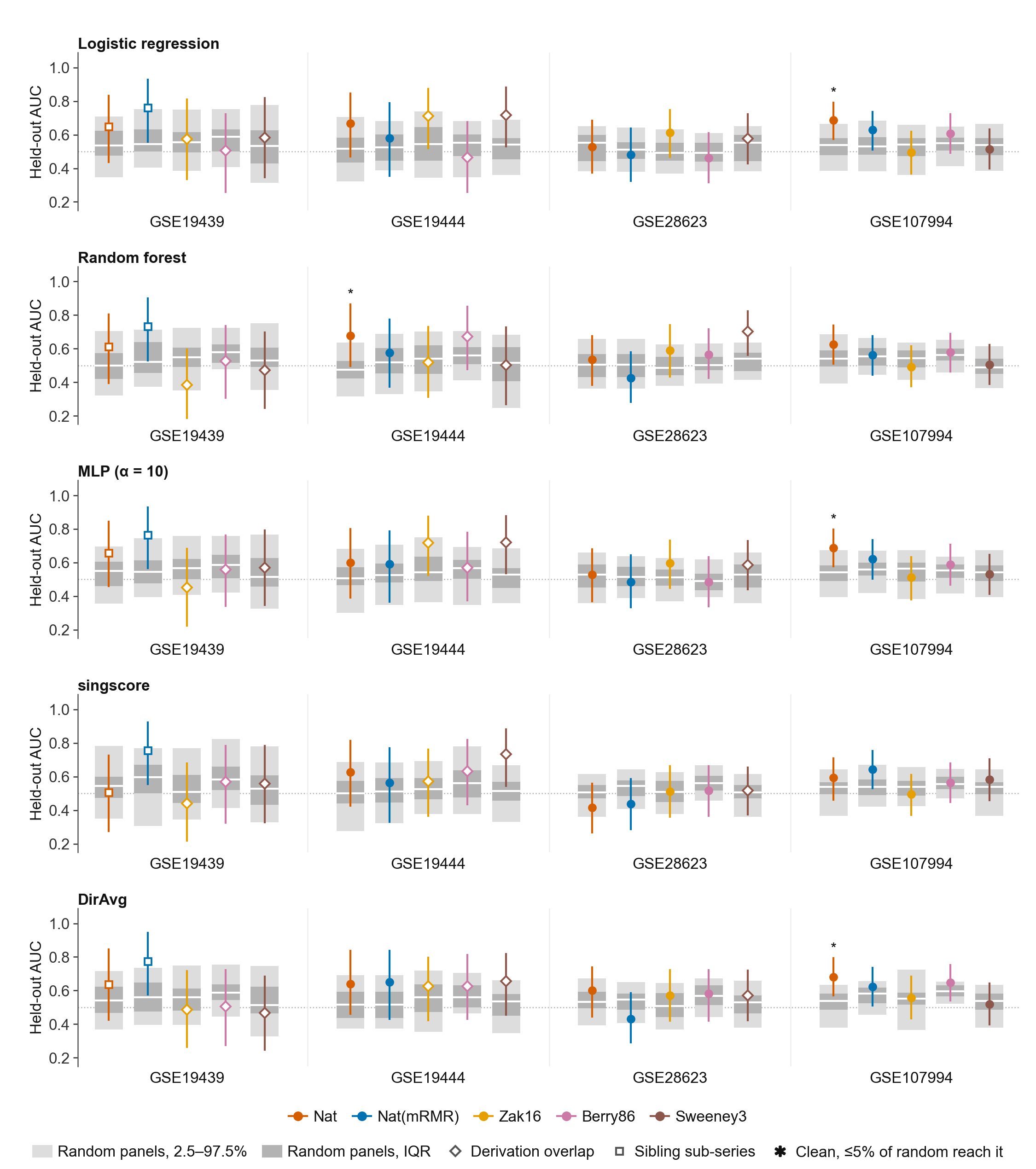

### S4 Fig. ATB vs LTB: derivation of Nat under all five scoring methods

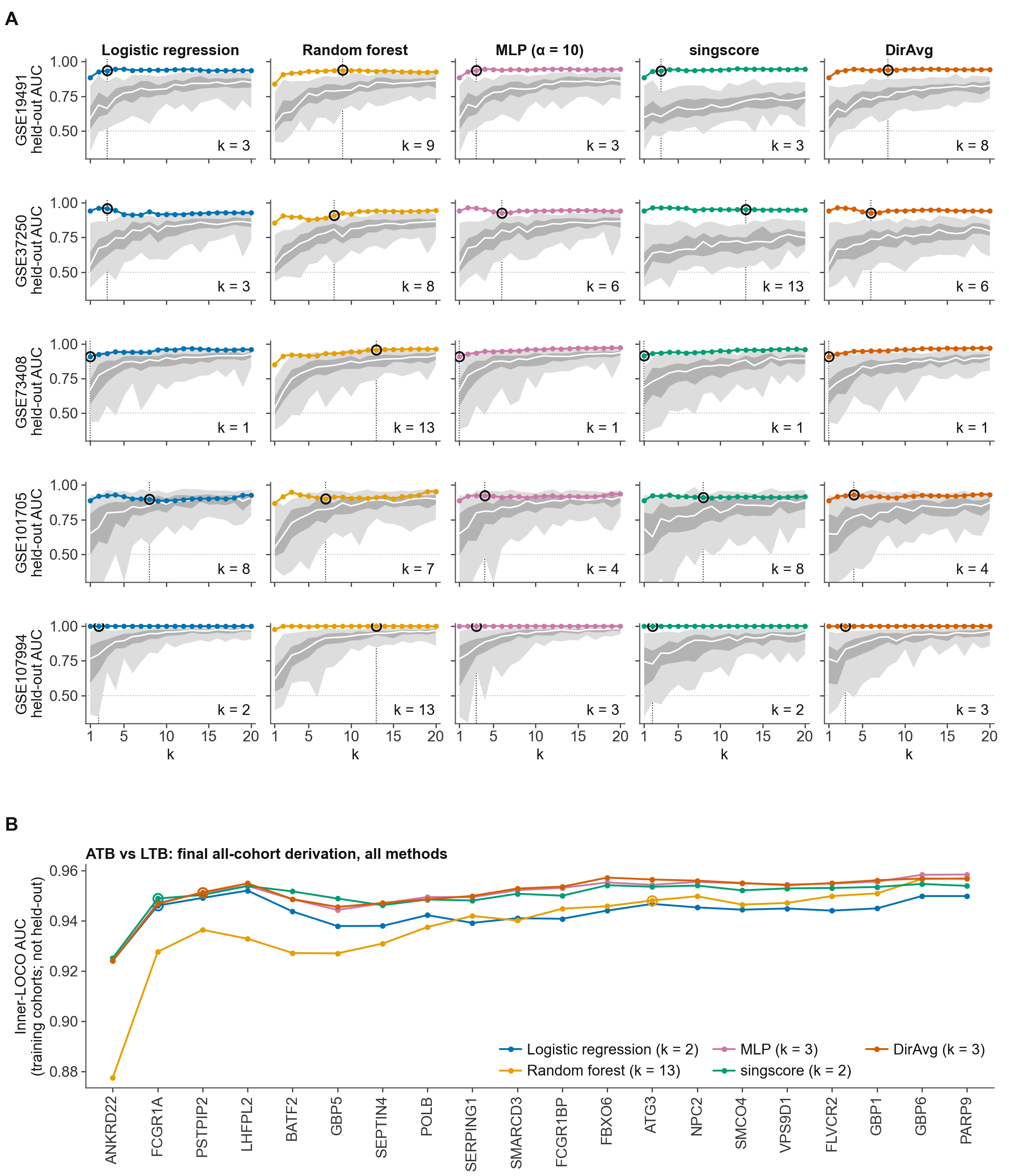

### S5 Fig. ATB vs OD: derivation of Nat under all five scoring methods

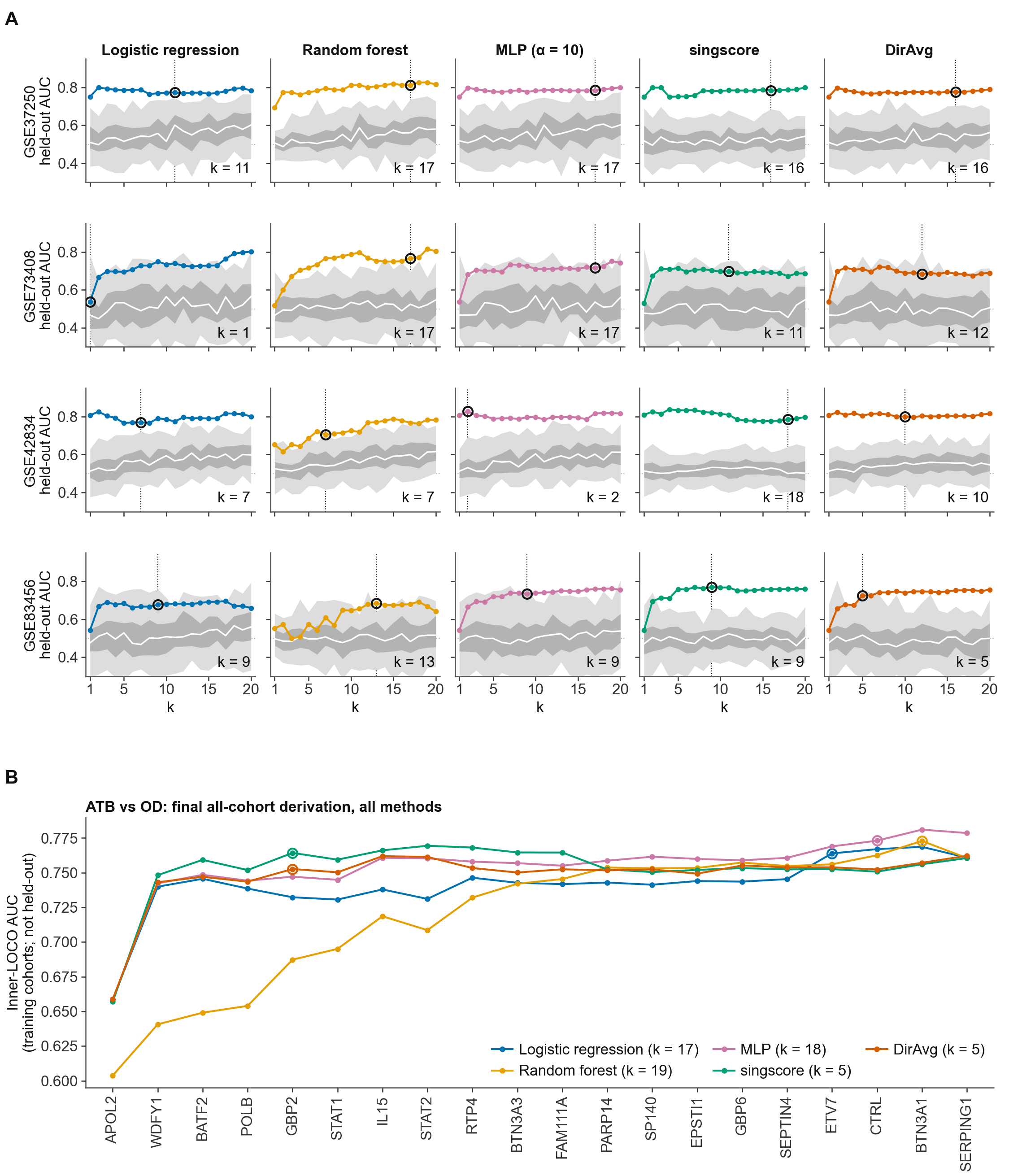

### S6 Fig. LTB vs CON: derivation of Nat under all five scoring methods

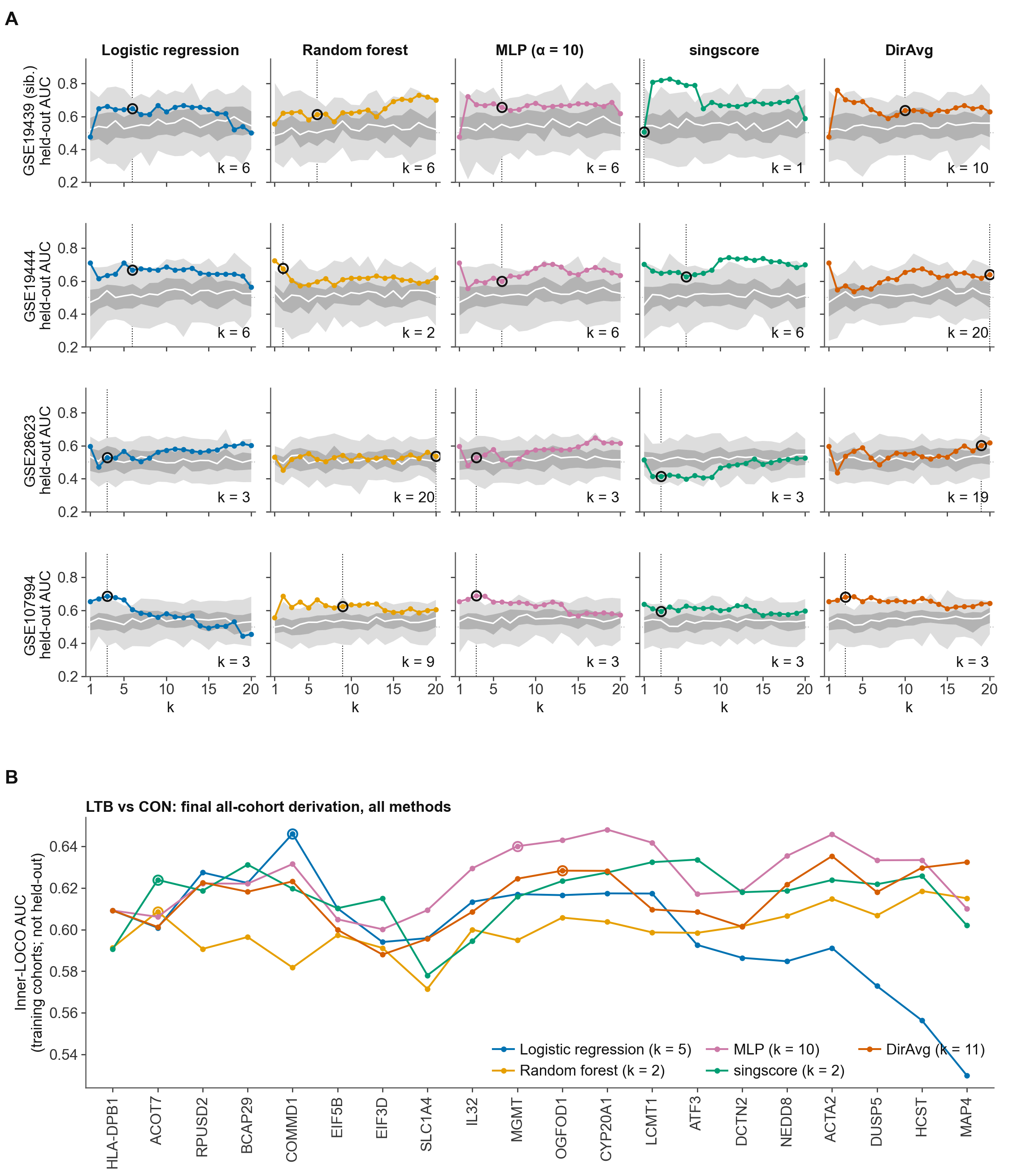

### S7 Fig. Every gene selected by the mRMR-derived panel, by fold

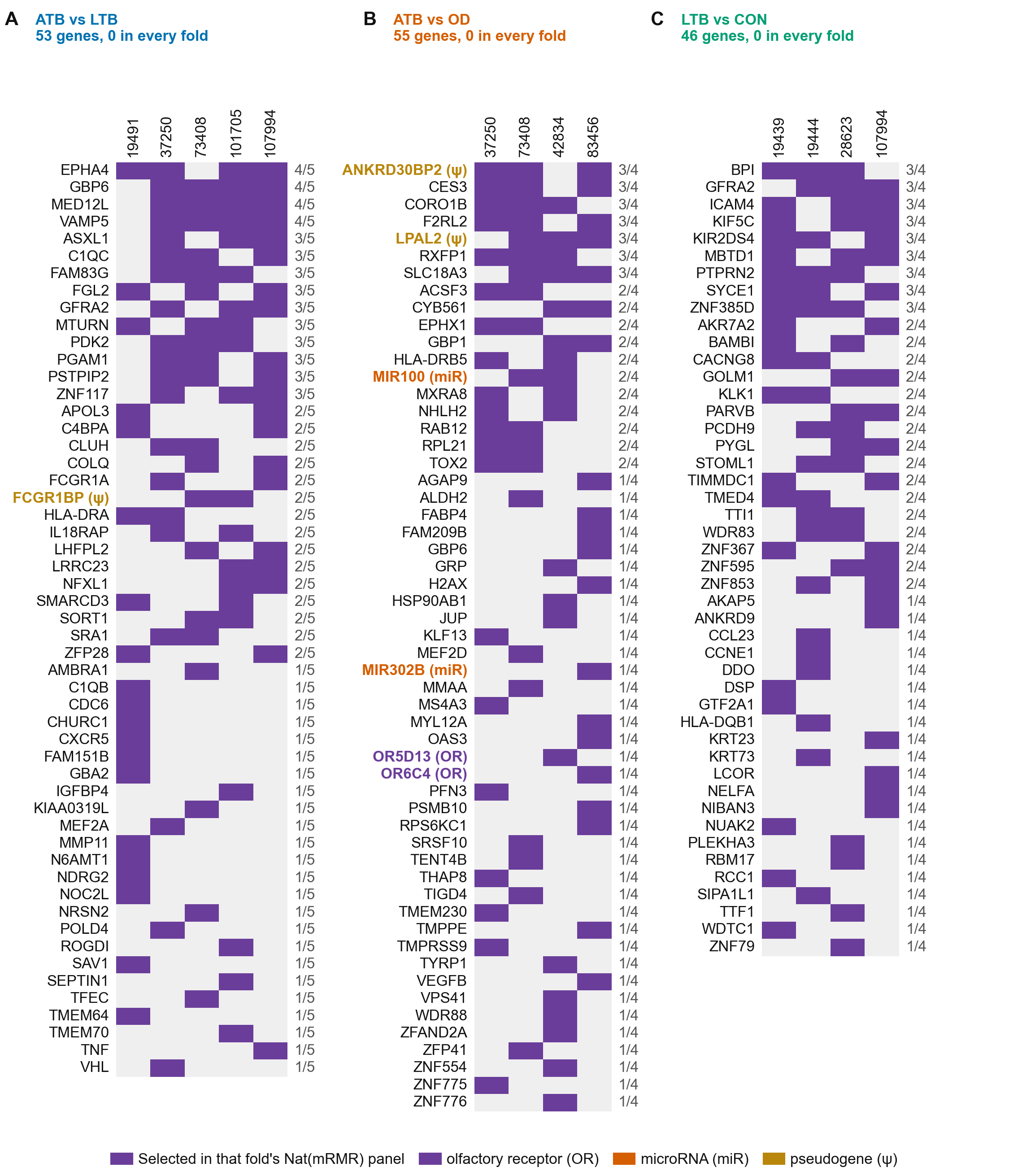

### S8 Fig. Cohort-level offset diagnostic

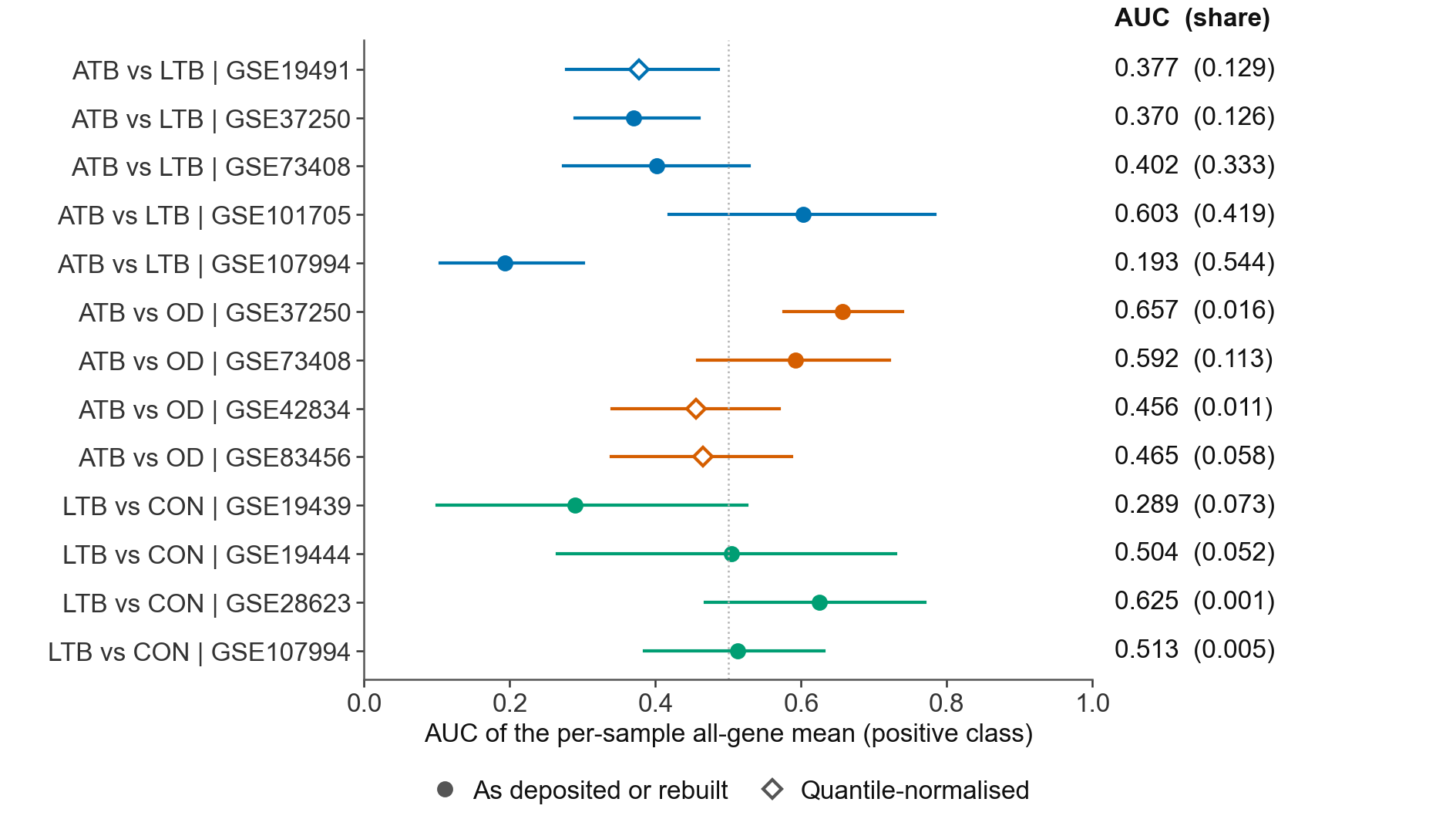
