## Supplementary material for "Cross-cohort benchmarking and validation reveals task-dependent generalisability in tuberculosis blood transcriptomic classification": S1 Table. ATB vs LTB: cross-cohort benchmark, all panels and scoring methods

**ATB vs LTB: cross-cohort benchmark, all panels and scoring methods.**

AUC in the held-out cohort for every panel and scoring method. Nat was rebuilt in each fold and its size chosen by an inner leave-one-cohort-out loop separately for each method, so the gene count is given as a range across the five methods; all other panels are fixed. The 95% confidence interval is given for the directional average, the primary score; intervals for the other methods are in the deposited analysis outputs. Random median is the median AUC of 50 size-matched random panels drawn for that fold and panel size. Beats random counts the scoring methods whose AUC was reached by at most 5% of those random panels. “Derivation overlap” marks cells where the published panel’s own derivation cohort, or one of its sub-series, is the held-out cohort; those AUCs are optimistic. Sibling sub-series marks the GSE19439 fold, whose training cohorts include its sister sub-series GSE19444. Values are not pooled or averaged across folds. Nat (per-fold) is not the same object as the frozen Nat (final) panel evaluated in S7-S10 Tables, which was derived once on all cohorts and therefore cannot be scored on any discovery cohort.

| Held-out cohort | Panel | Genes | LR | RF | MLP | singscore | DirAvg | DirAvg 95% CI | Random median | Beats random | Note |
| --- | --- | --- | --- | --- | --- | --- | --- | --- | --- | --- | --- |
| GSE101705 | Nat (per-fold) | 4 -8 | 0.897 | 0.901 | 0.924 | 0.911 | 0.929 | 0.844 -0.990 | 0.823 | 0/5 |  |
| GSE101705 | Nat(mRMR) | 20 | 0.929 | 0.921 | 0.926 | 0.935 | 0.915 | 0.831 -0.986 | 0.898 | 0/5 |  |
| GSE101705 | Berry86 | 73 | 0.971 | 0.958 | 0.973 | 0.955 | 0.955 | 0.891 -1.000 | 0.921 | 3/5 |  |
| GSE101705 | Kaforou (ATB vs LTB) | 22 | 0.911 | 0.945 | 0.933 | 0.915 | 0.938 | 0.855 -0.995 | 0.888 | 0/5 |  |
| GSE101705 | Sweeney3 | 3 | 0.904 | 0.907 | 0.895 | 0.920 | 0.837 | 0.720 -0.945 | 0.757 | 1/5 |  |
| GSE101705 | Zak16 | 16 | 0.953 | 0.944 | 0.926 | 0.933 | 0.922 | 0.830 -0.995 | 0.876 | 0/5 |  |
| GSE107994 | Nat (per-fold) | 2 -13 | 0.999 | 1.000 | 1.000 | 0.999 | 1.000 | 1.000 -1.000 | 0.834 | 5/5 |  |
| GSE107994 | Nat(mRMR) | 20 | 0.993 | 1.000 | 1.000 | 0.997 | 0.997 | 0.988 -1.000 | 0.968 | 4/5 |  |
| GSE107994 | Berry86 | 73 | 0.997 | 0.994 | 0.998 | 0.987 | 0.982 | 0.947 -1.000 | 0.984 | 1/5 |  |
| GSE107994 | Kaforou (ATB vs LTB) | 23 | 0.997 | 1.000 | 0.999 | 0.997 | 0.999 | 0.996 -1.000 | 0.972 | 4/5 |  |
| GSE107994 | Sweeney3 | 3 | 0.996 | 0.990 | 0.994 | 0.995 | 0.985 | 0.959 -0.999 | 0.834 | 5/5 |  |
| GSE107994 | Zak16 | 16 | 0.999 | 0.996 | 1.000 | 1.000 | 0.999 | 0.996 -1.000 | 0.963 | 5/5 |  |
| GSE19491 | Nat (per-fold) | 3 -9 | 0.935 | 0.938 | 0.937 | 0.934 | 0.941 | 0.889 -0.979 | 0.671 | 5/5 |  |
| GSE19491 | Nat(mRMR) | 20 | 0.913 | 0.907 | 0.924 | 0.852 | 0.897 | 0.827 -0.959 | 0.850 | 3/5 |  |
| GSE19491 | Berry86 | 73 | 0.902 | 0.934 | 0.939 | 0.949 | 0.947 | 0.899 -0.983 | 0.894 | 3/5 | Derivation overlap |
| GSE19491 | Kaforou (ATB vs LTB) | 22 | 0.944 | 0.949 | 0.957 | 0.954 | 0.955 | 0.910 -0.986 | 0.862 | 5/5 | Derivation overlap |
| GSE19491 | Sweeney3 | 3 | 0.919 | 0.935 | 0.921 | 0.928 | 0.914 | 0.848 -0.967 | 0.638 | 5/5 | Derivation overlap |
| GSE19491 | Zak16 | 16 | 0.922 | 0.930 | 0.937 | 0.938 | 0.935 | 0.876 -0.979 | 0.846 | 5/5 | Derivation overlap |
| GSE37250 | Nat (per-fold) | 3 -13 | 0.959 | 0.908 | 0.926 | 0.952 | 0.926 | 0.889 -0.958 | 0.738 | 5/5 |  |
| GSE37250 | Nat(mRMR) | 20 | 0.944 | 0.951 | 0.947 | 0.931 | 0.930 | 0.890 -0.966 | 0.856 | 5/5 |  |
| GSE37250 | Berry86 | 73 | 0.948 | 0.946 | 0.941 | 0.919 | 0.917 | 0.875 -0.956 | 0.883 | 4/5 |  |
| GSE37250 | Kaforou (ATB vs LTB) | 22 | 0.972 | 0.955 | 0.984 | 0.983 | 0.987 | 0.974 -0.996 | 0.855 | 5/5 | Derivation overlap |
| GSE37250 | Sweeney3 | 3 | 0.954 | 0.958 | 0.960 | 0.955 | 0.967 | 0.943 -0.987 | 0.664 | 5/5 | Derivation overlap |
| GSE37250 | Zak16 | 16 | 0.944 | 0.929 | 0.943 | 0.936 | 0.931 | 0.891 -0.967 | 0.841 | 5/5 | Derivation overlap |
| GSE73408 | Nat (per-fold) | 1 -13 | 0.909 | 0.957 | 0.909 | 0.916 | 0.909 | 0.827 -0.972 | 0.667 | 5/5 |  |
| GSE73408 | Nat(mRMR) | 20 | 0.949 | 0.934 | 0.962 | 0.971 | 0.974 | 0.939 -0.997 | 0.916 | 2/5 |  |
| GSE73408 | Berry86 | 74 | 0.931 | 0.980 | 0.980 | 0.980 | 0.980 | 0.952 -1.000 | 0.940 | 3/5 |  |
| GSE73408 | Kaforou (ATB vs LTB) | 22 | 0.964 | 0.971 | 0.980 | 0.980 | 0.981 | 0.952 -0.999 | 0.913 | 3/5 |  |
| GSE73408 | Sweeney3 | 3 | 0.967 | 0.925 | 0.966 | 0.973 | 0.947 | 0.891 -0.985 | 0.757 | 5/5 |  |
| GSE73408 | Zak16 | 16 | 0.921 | 0.927 | 0.938 | 0.936 | 0.939 | 0.876 -0.985 | 0.904 | 0/5 |  |
