## Supplementary material for "Cross-cohort benchmarking and validation reveals task-dependent generalisability in tuberculosis blood transcriptomic classification": S2 Table. ATB vs OD: cross-cohort benchmark, all panels and scoring methods

**ATB vs OD: cross-cohort benchmark, all panels and scoring methods.**

AUC in the held-out cohort for every panel and scoring method. Nat was rebuilt in each fold and its size chosen by an inner leave-one-cohort-out loop separately for each method, so the gene count is given as a range across the five methods; all other panels are fixed. The 95% confidence interval is given for the directional average, the primary score; intervals for the other methods are in the deposited analysis outputs. Random median is the median AUC of 50 size-matched random panels drawn for that fold and panel size. Beats random counts the scoring methods whose AUC was reached by at most 5% of those random panels. Derivation overlap marks cells where the published panel’s own derivation cohort, or one of its sub-series, is the held-out cohort; those AUCs are optimistic. Sibling sub-series marks the GSE19439 fold, whose training cohorts include its sister sub-series GSE19444. Values are not pooled or averaged across folds. Nat (per-fold) is not the same object as the frozen Nat (final) panel evaluated in S7-S10 Tables, which was derived once on all cohorts and therefore cannot be scored on any discovery cohort.

| Held-out cohort | Panel | Genes | LR | RF | MLP | singscore | DirAvg | DirAvg 95% CI | Random median | Beats random | Note |
| --- | --- | --- | --- | --- | --- | --- | --- | --- | --- | --- | --- |
| GSE37250 | Nat (per-fold) | 11 -17 | 0.772 | 0.811 | 0.784 | 0.783 | 0.775 | 0.706 -0.844 | 0.561 | 5/5 |  |
| GSE37250 | Nat(mRMR) | 20 | 0.650 | 0.682 | 0.656 | 0.457 | 0.551 | 0.465 -0.630 | 0.580 | 0/5 |  |
| GSE37250 | Berry86 | 73 | 0.750 | 0.792 | 0.790 | 0.646 | 0.704 | 0.626 -0.781 | 0.610 | 5/5 |  |
| GSE37250 | Kaforou (ATB vs OD) | 33 | 0.909 | 0.903 | 0.938 | 0.877 | 0.910 | 0.865 -0.953 | 0.613 | 5/5 | Derivation overlap |
| GSE37250 | Sweeney3 | 3 | 0.855 | 0.862 | 0.868 | 0.860 | 0.884 | 0.834 -0.932 | 0.526 | 5/5 | Derivation overlap |
| GSE37250 | Zak16 | 16 | 0.711 | 0.728 | 0.783 | 0.805 | 0.792 | 0.726 -0.859 | 0.558 | 5/5 | Derivation overlap |
| GSE42834 | Nat (per-fold) | 2 -18 | 0.769 | 0.704 | 0.827 | 0.786 | 0.800 | 0.712 -0.874 | 0.539 | 5/5 |  |
| GSE42834 | Nat(mRMR) | 20 | 0.616 | 0.683 | 0.629 | 0.447 | 0.584 | 0.469 -0.701 | 0.598 | 0/5 |  |
| GSE42834 | Berry86 | 73 | 0.796 | 0.880 | 0.844 | 0.619 | 0.826 | 0.740 -0.899 | 0.641 | 4/5 |  |
| GSE42834 | Kaforou (ATB vs OD) | 33 | 0.816 | 0.862 | 0.825 | 0.715 | 0.784 | 0.692 -0.867 | 0.646 | 5/5 |  |
| GSE42834 | Sweeney3 | 3 | 0.830 | 0.819 | 0.838 | 0.847 | 0.830 | 0.749 -0.899 | 0.513 | 5/5 | Derivation overlap |
| GSE42834 | Zak16 | 16 | 0.727 | 0.824 | 0.818 | 0.850 | 0.842 | 0.760 -0.908 | 0.580 | 4/5 | Derivation overlap |
| GSE73408 | Nat (per-fold) | 1 -17 | 0.537 | 0.766 | 0.716 | 0.698 | 0.684 | 0.557 -0.806 | 0.495 | 2/5 |  |
| GSE73408 | Nat(mRMR) | 20 | 0.399 | 0.428 | 0.407 | 0.626 | 0.544 | 0.419 -0.679 | 0.548 | 0/5 |  |
| GSE73408 | Berry86 | 74 | 0.760 | 0.837 | 0.812 | 0.607 | 0.661 | 0.527 -0.785 | 0.615 | 2/5 |  |
| GSE73408 | Kaforou (ATB vs OD) | 34 | 0.744 | 0.757 | 0.780 | 0.847 | 0.832 | 0.738 -0.917 | 0.549 | 4/5 |  |
| GSE73408 | Sweeney3 | 3 | 0.715 | 0.766 | 0.733 | 0.731 | 0.739 | 0.629 -0.848 | 0.480 | 5/5 |  |
| GSE73408 | Zak16 | 16 | 0.755 | 0.764 | 0.748 | 0.738 | 0.741 | 0.622 -0.851 | 0.486 | 5/5 |  |
| GSE83456 | Nat (per-fold) | 5 -13 | 0.677 | 0.684 | 0.733 | 0.770 | 0.725 | 0.619 -0.826 | 0.499 | 3/5 |  |
| GSE83456 | Nat(mRMR) | 20 | 0.481 | 0.490 | 0.512 | 0.547 | 0.599 | 0.480 -0.707 | 0.516 | 0/5 |  |
| GSE83456 | Berry86 | 73 | 0.732 | 0.580 | 0.754 | 0.533 | 0.653 | 0.542 -0.763 | 0.534 | 2/5 |  |
| GSE83456 | Kaforou (ATB vs OD) | 33 | 0.871 | 0.784 | 0.841 | 0.707 | 0.707 | 0.596 -0.802 | 0.504 | 5/5 |  |
| GSE83456 | Sweeney3 | 3 | 0.689 | 0.684 | 0.714 | 0.794 | 0.769 | 0.654 -0.870 | 0.486 | 3/5 |  |
| GSE83456 | Zak16 | 16 | 0.600 | 0.610 | 0.664 | 0.767 | 0.761 | 0.649 -0.858 | 0.523 | 2/5 |  |
