## Supplementary material for "Cross-cohort benchmarking and validation reveals task-dependent generalisability in tuberculosis blood transcriptomic classification": S3 Table. LTB vs CON: cross-cohort benchmark, all panels and scoring methods

**LTB vs CON: cross-cohort benchmark, all panels and scoring methods.**

AUC in the held-out cohort for every panel and scoring method. Nat was rebuilt in each fold and its size chosen by an inner leave-one-cohort-out loop separately for each method, so the gene count is given as a range across the five methods; all other panels are fixed. The 95% confidence interval is given for the directional average, the primary score; intervals for the other methods are in the deposited analysis outputs. Random median is the median AUC of 50 size-matched random panels drawn for that fold and panel size. Beats random counts the scoring methods whose AUC was reached by at most 5% of those random panels. Derivation overlap marks cells where the published panel’s own derivation cohort, or one of its sub-series, is the held-out cohort; those AUCs are optimistic. Sibling sub-series marks the GSE19439 fold, whose training cohorts include its sister sub-series GSE19444. Values are not pooled or averaged across folds. Nat (per-fold) is not the same object as the frozen Nat (final) panel evaluated in S7-S10 Tables, which was derived once on all cohorts and therefore cannot be scored on any discovery cohort.

| Held-out cohort | Panel | Genes | LR | RF | MLP | singscore | DirAvg | DirAvg 95% CI | Random median | Beats random | Note |
| --- | --- | --- | --- | --- | --- | --- | --- | --- | --- | --- | --- |
| GSE107994 | Nat (per-fold) | 3 -9 | 0.686 | 0.625 | 0.689 | 0.594 | 0.681 | 0.571 -0.793 | 0.540 | 3/5 |  |
| GSE107994 | Nat(mRMR) | 20 | 0.629 | 0.563 | 0.623 | 0.643 | 0.624 | 0.510 -0.736 | 0.553 | 0/5 |  |
| GSE107994 | Berry86 | 74 | 0.606 | 0.580 | 0.589 | 0.564 | 0.647 | 0.540 -0.751 | 0.557 | 0/5 |  |
| GSE107994 | Sweeney3 | 3 | 0.515 | 0.504 | 0.531 | 0.582 | 0.520 | 0.399 -0.641 | 0.539 | 0/5 |  |
| GSE107994 | Zak16 | 15 | 0.494 | 0.491 | 0.513 | 0.495 | 0.558 | 0.435 -0.683 | 0.543 | 0/5 |  |
| GSE19439 | Nat (per-fold) | 1 -10 | 0.647 | 0.613 | 0.657 | 0.507 | 0.637 | 0.426 -0.844 | 0.542 | 0/5 | Sibling sub-series |
| GSE19439 | Nat(mRMR) | 20 | 0.760 | 0.733 | 0.765 | 0.755 | 0.775 | 0.576 -0.944 | 0.544 | 5/5 | Sibling sub-series |
| GSE19439 | Berry86 | 74 | 0.505 | 0.527 | 0.559 | 0.569 | 0.505 | 0.274 -0.721 | 0.586 | 0/5 | Derivation overlap |
| GSE19439 | Sweeney3 | 3 | 0.583 | 0.473 | 0.569 | 0.559 | 0.466 | 0.248 -0.683 | 0.527 | 0/5 | Derivation overlap |
| GSE19439 | Zak16 | 15 | 0.574 | 0.385 | 0.451 | 0.441 | 0.485 | 0.263 -0.716 | 0.554 | 0/5 | Derivation overlap |
| GSE19444 | Nat (per-fold) | 2 -20 | 0.667 | 0.677 | 0.599 | 0.627 | 0.639 | 0.462 -0.837 | 0.508 | 1/5 |  |
| GSE19444 | Nat(mRMR) | 20 | 0.579 | 0.575 | 0.591 | 0.563 | 0.651 | 0.431 -0.838 | 0.518 | 0/5 |  |
| GSE19444 | Berry86 | 74 | 0.464 | 0.671 | 0.571 | 0.631 | 0.627 | 0.431 -0.812 | 0.560 | 0/5 | Derivation overlap |
| GSE19444 | Sweeney3 | 3 | 0.718 | 0.502 | 0.722 | 0.734 | 0.655 | 0.456 -0.817 | 0.528 | 4/5 | Derivation overlap |
| GSE19444 | Zak16 | 15 | 0.710 | 0.518 | 0.718 | 0.571 | 0.627 | 0.422 -0.796 | 0.542 | 0/5 | Derivation overlap |
| GSE28623 | Nat (per-fold) | 3 -20 | 0.528 | 0.536 | 0.529 | 0.415 | 0.602 | 0.446 -0.739 | 0.528 | 0/5 |  |
| GSE28623 | Nat(mRMR) | 20 | 0.481 | 0.425 | 0.484 | 0.437 | 0.432 | 0.291 -0.585 | 0.515 | 0/5 |  |
| GSE28623 | Berry86 | 74 | 0.463 | 0.564 | 0.486 | 0.516 | 0.581 | 0.420 -0.722 | 0.503 | 0/5 |  |
| GSE28623 | Sweeney3 | 3 | 0.578 | 0.702 | 0.587 | 0.516 | 0.572 | 0.422 -0.718 | 0.529 | 1/5 | Derivation overlap |
| GSE28623 | Zak16 | 16 | 0.612 | 0.589 | 0.596 | 0.512 | 0.572 | 0.420 -0.722 | 0.508 | 0/5 |  |
