## Supplementary material for "Cross-cohort benchmarking and validation reveals task-dependent generalisability in tuberculosis blood transcriptomic classification": S4 Table. Nat panel size selected in each fold by the inner leave-one-cohort-out loop

**Nat panel size selected in each fold by the inner leave-one-cohort-out loop.**

For each fold the direction-consistency ranking was rebuilt on the training cohorts only, and k (1-20) was chosen as the smallest value whose mean inner-loop AUC was within 0.01 of the best, separately for each scoring method. Within a fold the ranking is identical for all scoring methods, which differ only in where it is cut; the panel selected in each fold is the first k genes of that fold’s ranking, and the full per-fold rankings are in the deposited analysis outputs. The final column is the held-out AUC at the chosen k minus the held-out AUC a fixed 20-gene panel would have given in the same fold.

| Task | Held-out cohort | Method | Chosen k | AUC at chosen k | 95% CI | Random median | Prop. random ≥ | Chosen - k=20 |
| --- | --- | --- | --- | --- | --- | --- | --- | --- |
| ATB vs LTB | GSE101705 | LR | 8 | 0.897 | 0.785 -0.983 | 0.850 | 0.26 | -0.029 |
| ATB vs LTB | GSE101705 | RF | 7 | 0.901 | 0.786 -0.992 | 0.823 | 0.14 | -0.052 |
| ATB vs LTB | GSE101705 | MLP | 4 | 0.924 | 0.834 -0.990 | 0.811 | 0.06 | -0.011 |
| ATB vs LTB | GSE101705 | singscore | 8 | 0.911 | 0.815 -0.985 | 0.824 | 0.06 | -0.004 |
| ATB vs LTB | GSE101705 | DirAvg | 4 | 0.929 | 0.844 -0.990 | 0.772 | 0.08 | -0.002 |
| ATB vs LTB | GSE107994 | LR | 2 | 0.999 | 0.993 -1.000 | 0.794 | 0.00 | -0.001 |
| ATB vs LTB | GSE107994 | RF | 13 | 1.000 | 0.998 -1.000 | 0.948 | 0.00 | +0.001 |
| ATB vs LTB | GSE107994 | MLP | 3 | 1.000 | 1.000 -1.000 | 0.847 | 0.00 | +0.000 |
| ATB vs LTB | GSE107994 | singscore | 2 | 0.999 | 0.993 -1.000 | 0.726 | 0.00 | -0.001 |
| ATB vs LTB | GSE107994 | DirAvg | 3 | 1.000 | 1.000 -1.000 | 0.834 | 0.00 | +0.000 |
| ATB vs LTB | GSE19491 | LR | 3 | 0.935 | 0.881 -0.975 | 0.662 | 0.00 | -0.002 |
| ATB vs LTB | GSE19491 | RF | 9 | 0.938 | 0.885 -0.977 | 0.793 | 0.00 | +0.012 |
| ATB vs LTB | GSE19491 | MLP | 3 | 0.937 | 0.882 -0.976 | 0.671 | 0.00 | -0.009 |
| ATB vs LTB | GSE19491 | singscore | 3 | 0.934 | 0.877 -0.975 | 0.604 | 0.00 | -0.013 |
| ATB vs LTB | GSE19491 | DirAvg | 8 | 0.941 | 0.889 -0.979 | 0.743 | 0.00 | -0.003 |
| ATB vs LTB | GSE37250 | LR | 3 | 0.959 | 0.932 -0.982 | 0.691 | 0.00 | +0.031 |
| ATB vs LTB | GSE37250 | RF | 8 | 0.908 | 0.861 -0.949 | 0.809 | 0.04 | -0.036 |
| ATB vs LTB | GSE37250 | MLP | 6 | 0.926 | 0.888 -0.958 | 0.803 | 0.00 | -0.014 |
| ATB vs LTB | GSE37250 | singscore | 13 | 0.952 | 0.919 -0.980 | 0.707 | 0.00 | +0.003 |
| ATB vs LTB | GSE37250 | DirAvg | 6 | 0.926 | 0.889 -0.958 | 0.738 | 0.00 | -0.014 |
| ATB vs LTB | GSE73408 | LR | 1 | 0.909 | 0.827 -0.972 | 0.667 | 0.02 | -0.051 |
| ATB vs LTB | GSE73408 | RF | 13 | 0.957 | 0.898 -0.993 | 0.883 | 0.00 | -0.007 |
| ATB vs LTB | GSE73408 | MLP | 1 | 0.909 | 0.827 -0.972 | 0.664 | 0.02 | -0.063 |
| ATB vs LTB | GSE73408 | singscore | 1 | 0.916 | 0.840 -0.978 | 0.686 | 0.02 | -0.042 |
| ATB vs LTB | GSE73408 | DirAvg | 1 | 0.909 | 0.827 -0.972 | 0.667 | 0.02 | -0.060 |
| ATB vs OD | GSE37250 | LR | 11 | 0.772 | 0.705 -0.843 | 0.601 | 0.02 | -0.010 |
| ATB vs OD | GSE37250 | RF | 17 | 0.811 | 0.749 -0.871 | 0.561 | 0.00 | -0.004 |
| ATB vs OD | GSE37250 | MLP | 17 | 0.784 | 0.716 -0.849 | 0.598 | 0.00 | -0.016 |
| ATB vs OD | GSE37250 | singscore | 16 | 0.783 | 0.716 -0.849 | 0.510 | 0.00 | -0.016 |
| ATB vs OD | GSE37250 | DirAvg | 16 | 0.775 | 0.706 -0.844 | 0.518 | 0.00 | -0.014 |
| ATB vs OD | GSE42834 | LR | 7 | 0.769 | 0.672 -0.859 | 0.571 | 0.00 | -0.031 |
| ATB vs OD | GSE42834 | RF | 7 | 0.704 | 0.600 -0.795 | 0.539 | 0.04 | -0.078 |
| ATB vs OD | GSE42834 | MLP | 2 | 0.827 | 0.733 -0.900 | 0.531 | 0.00 | +0.010 |
| ATB vs OD | GSE42834 | singscore | 18 | 0.786 | 0.696 -0.871 | 0.500 | 0.00 | -0.012 |
| ATB vs OD | GSE42834 | DirAvg | 10 | 0.800 | 0.712 -0.874 | 0.557 | 0.00 | -0.015 |
| ATB vs OD | GSE73408 | LR | 1 | 0.537 | 0.398 -0.668 | 0.468 | 0.28 | -0.264 |
| ATB vs OD | GSE73408 | RF | 17 | 0.766 | 0.652 -0.868 | 0.525 | 0.02 | -0.038 |
| ATB vs OD | GSE73408 | MLP | 17 | 0.716 | 0.596 -0.834 | 0.536 | 0.02 | -0.028 |
| ATB vs OD | GSE73408 | singscore | 11 | 0.698 | 0.568 -0.819 | 0.493 | 0.08 | +0.012 |
| ATB vs OD | GSE73408 | DirAvg | 12 | 0.684 | 0.557 -0.806 | 0.495 | 0.10 | -0.006 |
| ATB vs OD | GSE83456 | LR | 9 | 0.677 | 0.561 -0.785 | 0.499 | 0.12 | +0.018 |
| ATB vs OD | GSE83456 | RF | 13 | 0.684 | 0.560 -0.794 | 0.502 | 0.00 | +0.043 |
| ATB vs OD | GSE83456 | MLP | 9 | 0.733 | 0.622 -0.828 | 0.480 | 0.04 | -0.021 |
| ATB vs OD | GSE83456 | singscore | 9 | 0.770 | 0.662 -0.863 | 0.485 | 0.00 | +0.010 |
| ATB vs OD | GSE83456 | DirAvg | 5 | 0.725 | 0.619 -0.826 | 0.516 | 0.06 | -0.030 |
| LTB vs CON | GSE107994 | LR | 3 | 0.686 | 0.574 -0.792 | 0.540 | 0.02 | +0.231 |
| LTB vs CON | GSE107994 | RF | 9 | 0.625 | 0.509 -0.737 | 0.541 | 0.20 | +0.021 |
| LTB vs CON | GSE107994 | MLP | 3 | 0.689 | 0.578 -0.797 | 0.544 | 0.02 | +0.118 |
| LTB vs CON | GSE107994 | singscore | 3 | 0.594 | 0.463 -0.708 | 0.539 | 0.12 | -0.003 |
| LTB vs CON | GSE107994 | DirAvg | 3 | 0.681 | 0.571 -0.793 | 0.538 | 0.00 | +0.037 |
| LTB vs CON | GSE19439 | LR | 6 | 0.647 | 0.438 -0.832 | 0.537 | 0.16 | +0.147 |
| LTB vs CON | GSE19439 | RF | 6 | 0.613 | 0.394 -0.803 | 0.500 | 0.14 | -0.086 |
| LTB vs CON | GSE19439 | MLP | 6 | 0.657 | 0.461 -0.843 | 0.551 | 0.14 | +0.039 |
| LTB vs CON | GSE19439 | singscore | 1 | 0.507 | 0.277 -0.726 | 0.544 | 0.52 | -0.081 |
| LTB vs CON | GSE19439 | DirAvg | 10 | 0.637 | 0.426 -0.844 | 0.542 | 0.22 | +0.010 |
| LTB vs CON | GSE19444 | LR | 6 | 0.667 | 0.471 -0.845 | 0.518 | 0.12 | +0.103 |
| LTB vs CON | GSE19444 | RF | 2 | 0.677 | 0.496 -0.865 | 0.475 | 0.02 | +0.056 |
| LTB vs CON | GSE19444 | MLP | 6 | 0.599 | 0.393 -0.801 | 0.508 | 0.16 | -0.036 |
| LTB vs CON | GSE19444 | singscore | 6 | 0.627 | 0.427 -0.814 | 0.500 | 0.10 | -0.071 |
| LTB vs CON | GSE19444 | DirAvg | 20 | 0.639 | 0.462 -0.837 | 0.516 | 0.10 | +0.000 |
| LTB vs CON | GSE28623 | LR | 3 | 0.528 | 0.373 -0.683 | 0.552 | 0.54 | -0.074 |
| LTB vs CON | GSE28623 | RF | 20 | 0.536 | 0.384 -0.676 | 0.505 | 0.34 | +0.000 |
| LTB vs CON | GSE28623 | MLP | 3 | 0.529 | 0.369 -0.679 | 0.528 | 0.50 | -0.088 |
| LTB vs CON | GSE28623 | singscore | 3 | 0.415 | 0.268 -0.559 | 0.505 | 0.96 | -0.109 |
| LTB vs CON | GSE28623 | DirAvg | 19 | 0.602 | 0.446 -0.739 | 0.531 | 0.18 | -0.017 |
