## Supplementary material for "Cross-cohort benchmarking and validation reveals task-dependent generalisability in tuberculosis blood transcriptomic classification": S5 Table. Comparison of six feature-selection criteria

**Comparison of six feature-selection criteria.**

Every variant builds a 20-gene panel per fold, is rebuilt inside the fold using the training cohorts only, and is scored once on the held-out cohort. Median, minimum and maximum are taken over all fold-by-method cells of the task (25 for ATB vs LTB, 20 for the other two). Beats random counts the cells whose AUC was reached by at most 5% of 50 size-matched random panels. Note that these are fixed 20-gene panels, so the Nat row here is not identical to the inner-loop-sized Nat reported in S1-S4 Tables.

| Task | Selection criterion | Median AUC | Min | Max | Beats random |
| --- | --- | --- | --- | --- | --- |
| ATB vs LTB | Nat(mRMR), as specified | 0.935 | 0.852 | 1.000 | 14/25 |
| ATB vs LTB | mRMR, relevance and redundancy rescaled | 0.951 | 0.884 | 1.000 | 19/25 |
| ATB vs LTB | MI top-20 union + forward selection | 0.954 | 0.920 | 0.999 | 19/25 |
| ATB vs LTB | MI mean rank | 0.960 | 0.913 | 1.000 | 19/25 |
| ATB vs LTB | Direction consistency + forward selection | 0.960 | 0.917 | 0.999 | 19/25 |
| ATB vs LTB | Direction consistency (Nat) | 0.947 | 0.915 | 1.000 | 17/25 |
| ATB vs OD | Nat(mRMR), as specified | 0.549 | 0.399 | 0.683 | 0/20 |
| ATB vs OD | mRMR, relevance and redundancy rescaled | 0.613 | 0.484 | 0.717 | 1/20 |
| ATB vs OD | MI top-20 union + forward selection | 0.712 | 0.524 | 0.814 | 7/20 |
| ATB vs OD | MI mean rank | 0.722 | 0.612 | 0.824 | 10/20 |
| ATB vs OD | Direction consistency + forward selection | 0.770 | 0.632 | 0.827 | 13/20 |
| ATB vs OD | Direction consistency (Nat) | 0.786 | 0.641 | 0.817 | 13/20 |
| LTB vs CON | Nat(mRMR), as specified | 0.607 | 0.425 | 0.775 | 5/20 |
| LTB vs CON | mRMR, relevance and redundancy rescaled | 0.573 | 0.444 | 0.711 | 0/20 |
| LTB vs CON | MI top-20 union + forward selection | 0.550 | 0.470 | 0.746 | 2/20 |
| LTB vs CON | MI mean rank | 0.549 | 0.438 | 0.701 | 1/20 |
| LTB vs CON | Direction consistency + forward selection | 0.564 | 0.431 | 0.667 | 0/20 |
| LTB vs CON | Direction consistency (Nat) | 0.610 | 0.455 | 0.699 | 1/20 |
