## Supplementary material for "Cross-cohort benchmarking and validation reveals task-dependent generalisability in tuberculosis blood transcriptomic classification": S7 Table. External validation: locked panels applied without refitting

**External validation: locked panels applied without refitting.**

GSE144127 (ATB vs OD; 79 active TB vs 300 other disease) and GSE152218 (ATB vs LTB; 14 vs 27). Neither cohort took part in panel derivation, model training or any earlier analysis. Models were trained on all discovery cohorts of the task and scored once. Nat (final) is the frozen panel for that task, derived once on all discovery cohorts of the task and applied here without modification: ANKRD22, FCGR1A and PSTPIP2 for ATB vs LTB (k = 3) and APOL2, WDFY1, BATF2, POLB and GBP2 for ATB vs OD (k = 5). Combined8 is the union of those two panels, an eight-gene set applied unchanged to both tasks. MI mean rank is the corresponding frozen panel from the MI mean-rank selector variant. Random median and the empirical p-value compare the panel with 50 size-matched random panels drawn from the same gene universe. These were drawn only for the panels derived in this study, since the question they address is whether our own selection beats chance; the published panels are compared with one another in S8 Table instead. Empirical p is (number of random panels reaching the observed AUC + 1) / 51, so 0.020 is the smallest value obtainable with 50 draws and is a floor rather than a point estimate. These p-values are not adjusted for multiplicity and should be read across the five scoring methods of a panel together, not singly. The first half of the table represents ATB v OD external testing (GSE 144127), the second half represents ATB v LTB external testing (GSE 152218)

| Cohort | Panel | Genes | Method | AUC | 95% CI | Random median | Empirical p |
| --- | --- | --- | --- | --- | --- | --- | --- |
| GSE144127 | Nat (final) | 5 | LR | 0.848 | 0.798 -0.892 | 0.530 | 0.020 |
| GSE144127 | Nat (final) | 5 | RF | 0.734 | 0.675 -0.782 | 0.510 | 0.020 |
| GSE144127 | Nat (final) | 5 | MLP | 0.853 | 0.803 -0.896 | 0.533 | 0.020 |
| GSE144127 | Nat (final) | 5 | singscore | 0.861 | 0.813 -0.902 | 0.514 | 0.020 |
| GSE144127 | Nat (final) | 5 | DirAvg | 0.855 | 0.806 -0.898 | 0.509 | 0.020 |
| GSE144127 | Combined8 | 8 | LR | 0.843 | 0.786 -0.888 | 0.542 | 0.020 |
| GSE144127 | Combined8 | 8 | RF | 0.784 | 0.729 -0.829 | 0.537 | 0.020 |
| GSE144127 | Combined8 | 8 | MLP | 0.857 | 0.809 -0.899 | 0.559 | 0.020 |
| GSE144127 | Combined8 | 8 | singscore | 0.860 | 0.811 -0.901 | 0.532 | 0.020 |
| GSE144127 | Combined8 | 8 | DirAvg | 0.858 | 0.808 -0.900 | 0.526 | 0.020 |
| GSE144127 | MI mean rank | 4 | LR | 0.824 | 0.768 -0.871 | 0.538 | 0.020 |
| GSE144127 | MI mean rank | 4 | RF | 0.732 | 0.669 -0.788 | 0.522 | 0.039 |
| GSE144127 | MI mean rank | 4 | MLP | 0.836 | 0.784 -0.880 | 0.526 | 0.020 |
| GSE144127 | MI mean rank | 4 | singscore | 0.746 | 0.691 -0.797 | 0.519 | 0.020 |
| GSE144127 | MI mean rank | 4 | DirAvg | 0.825 | 0.775 -0.870 | 0.498 | 0.020 |
| GSE144127 | Berry86 | 74 | LR | 0.693 | 0.622 -0.761 |  |  |
| GSE144127 | Berry86 | 74 | RF | 0.824 | 0.768 -0.872 |  |  |
| GSE144127 | Berry86 | 74 | MLP | 0.784 | 0.720 -0.842 |  |  |
| GSE144127 | Berry86 | 74 | singscore | 0.740 | 0.675 -0.798 |  |  |
| GSE144127 | Berry86 | 74 | DirAvg | 0.839 | 0.784 -0.887 |  |  |
| GSE144127 | Kaforou (ATB vs OD) | 34 | LR | 0.751 | 0.677 -0.813 |  |  |
| GSE144127 | Kaforou (ATB vs OD) | 34 | RF | 0.777 | 0.712 -0.835 |  |  |
| GSE144127 | Kaforou (ATB vs OD) | 34 | MLP | 0.781 | 0.711 -0.839 |  |  |
| GSE144127 | Kaforou (ATB vs OD) | 34 | singscore | 0.794 | 0.736 -0.847 |  |  |
| GSE144127 | Kaforou (ATB vs OD) | 34 | DirAvg | 0.821 | 0.767 -0.867 |  |  |
| GSE144127 | Sweeney3 | 3 | LR | 0.827 | 0.767 -0.879 |  |  |
| GSE144127 | Sweeney3 | 3 | RF | 0.767 | 0.708 -0.826 |  |  |
| GSE144127 | Sweeney3 | 3 | MLP | 0.824 | 0.763 -0.878 |  |  |
| GSE144127 | Sweeney3 | 3 | singscore | 0.853 | 0.802 -0.900 |  |  |
| GSE144127 | Sweeney3 | 3 | DirAvg | 0.808 | 0.744 -0.864 |  |  |
| GSE144127 | Zak16 | 16 | LR | 0.856 | 0.807 -0.898 |  |  |
| GSE144127 | Zak16 | 16 | RF | 0.832 | 0.782 -0.873 |  |  |
| GSE144127 | Zak16 | 16 | MLP | 0.865 | 0.820 -0.905 |  |  |
| GSE144127 | Zak16 | 16 | singscore | 0.863 | 0.818 -0.902 |  |  |
| GSE144127 | Zak16 | 16 | DirAvg | 0.859 | 0.811 -0.899 |  |  |
| Cohort | Panel | Genes | Method | AUC | 95% CI | Random median | Empirical p |
| GSE152218 | Nat (final) | 3 | LR | 0.963 | 0.898 -1.000 | 0.886 | 0.176 |
| GSE152218 | Nat (final) | 3 | RF | 0.927 | 0.836 -0.992 | 0.837 | 0.157 |
| GSE152218 | Nat (final) | 3 | MLP | 0.960 | 0.892 -1.000 | 0.878 | 0.176 |
| GSE152218 | Nat (final) | 3 | singscore | 0.955 | 0.881 -1.000 | 0.806 | 0.078 |
| GSE152218 | Nat (final) | 3 | DirAvg | 0.963 | 0.903 -1.000 | 0.825 | 0.039 |
| GSE152218 | Combined8 | 8 | LR | 0.960 | 0.897 -1.000 | 0.956 | 0.451 |
| GSE152218 | Combined8 | 8 | RF | 0.960 | 0.900 -1.000 | 0.931 | 0.216 |
| GSE152218 | Combined8 | 8 | MLP | 0.955 | 0.883 -1.000 | 0.955 | 0.549 |
| GSE152218 | Combined8 | 8 | singscore | 0.942 | 0.862 -0.991 | 0.910 | 0.333 |
| GSE152218 | Combined8 | 8 | DirAvg | 0.947 | 0.873 -0.998 | 0.931 | 0.353 |
| GSE152218 | MI mean rank | 3 | LR | 0.936 | 0.860 -0.988 | 0.886 | 0.275 |
| GSE152218 | MI mean rank | 3 | RF | 0.914 | 0.816 -0.981 | 0.837 | 0.216 |
| GSE152218 | MI mean rank | 3 | MLP | 0.944 | 0.874 -0.991 | 0.878 | 0.176 |
| GSE152218 | MI mean rank | 3 | singscore | 0.939 | 0.867 -0.988 | 0.806 | 0.137 |
| GSE152218 | MI mean rank | 3 | DirAvg | 0.944 | 0.875 -0.991 | 0.825 | 0.118 |
| GSE152218 | Berry86 | 74 | LR | 0.997 | 0.984 -1.000 |  |  |
| GSE152218 | Berry86 | 74 | RF | 0.984 | 0.952 -1.000 |  |  |
| GSE152218 | Berry86 | 74 | MLP | 1.000 | 1.000 -1.000 |  |  |
| GSE152218 | Berry86 | 74 | singscore | 0.987 | 0.954 -1.000 |  |  |
| GSE152218 | Berry86 | 74 | DirAvg | 0.989 | 0.962 -1.000 |  |  |
| GSE152218 | Kaforou (ATB vs LTB) | 23 | LR | 0.958 | 0.884 -1.000 |  |  |
| GSE152218 | Kaforou (ATB vs LTB) | 23 | RF | 0.955 | 0.870 -1.000 |  |  |
| GSE152218 | Kaforou (ATB vs LTB) | 23 | MLP | 0.960 | 0.891 -1.000 |  |  |
| GSE152218 | Kaforou (ATB vs LTB) | 23 | singscore | 0.966 | 0.899 -1.000 |  |  |
| GSE152218 | Kaforou (ATB vs LTB) | 23 | DirAvg | 0.963 | 0.896 -1.000 |  |  |
| GSE152218 | Sweeney3 | 3 | LR | 0.960 | 0.898 -1.000 |  |  |
| GSE152218 | Sweeney3 | 3 | RF | 0.960 | 0.893 -1.000 |  |  |
| GSE152218 | Sweeney3 | 3 | MLP | 0.960 | 0.898 -1.000 |  |  |
| GSE152218 | Sweeney3 | 3 | singscore | 0.976 | 0.929 -1.000 |  |  |
| GSE152218 | Sweeney3 | 3 | DirAvg | 0.944 | 0.876 -0.991 |  |  |
| GSE152218 | Zak16 | 16 | LR | 0.950 | 0.865 -1.000 |  |  |
| GSE152218 | Zak16 | 16 | RF | 0.911 | 0.754 -0.998 |  |  |
| GSE152218 | Zak16 | 16 | MLP | 0.944 | 0.849 -1.000 |  |  |
| GSE152218 | Zak16 | 16 | singscore | 0.915 | 0.815 -0.984 |  |  |
| GSE152218 | Zak16 | 16 | DirAvg | 0.926 | 0.824 -0.992 |  |  |
