## Supplementary material for "Cross-cohort benchmarking and validation reveals task-dependent generalisability in tuberculosis blood transcriptomic classification": S8 Table. Paired DeLong comparisons in the external cohorts

**Paired DeLong comparisons in the external cohorts.**

Nat (final) against every other panel, computed on identical samples. ΔAUC is Nat (final) minus comparator. p-values are adjusted by the Holm procedure within each cohort and scoring method, over the six comparators listed. No comparison reaches significance after adjustment in either cohort.

| Cohort | Method | Comparator | AUC Nat (final) | AUC comparator | ΔAUC | 95% CI | p | p (Holm) |
| --- | --- | --- | --- | --- | --- | --- | --- | --- |
| GSE144127 | LR | Berry86 | 0.848 | 0.693 | +0.1548 | +0.0910 to +0.2186 | 0.000 | 0.000 |
| GSE144127 | LR | Combined8 | 0.848 | 0.843 | +0.0048 | -0.0217 to +0.0313 | 0.722 | 1.000 |
| GSE144127 | LR | Kaforou (ATB vs OD) | 0.848 | 0.751 | +0.0968 | +0.0412 to +0.1524 | 0.001 | 0.003 |
| GSE144127 | LR | MI mean rank | 0.848 | 0.824 | +0.0240 | -0.0067 to +0.0547 | 0.125 | 0.500 |
| GSE144127 | LR | Sweeney3 | 0.848 | 0.827 | +0.0216 | -0.0181 to +0.0614 | 0.286 | 0.857 |
| GSE144127 | LR | Zak16 | 0.848 | 0.856 | -0.0081 | -0.0479 to +0.0317 | 0.690 | 1.000 |
| GSE144127 | RF | Berry86 | 0.734 | 0.824 | -0.0900 | -0.1396 to -0.0405 | 0.000 | 0.002 |
| GSE144127 | RF | Combined8 | 0.734 | 0.784 | -0.0503 | -0.0790 to -0.0217 | 0.001 | 0.002 |
| GSE144127 | RF | Kaforou (ATB vs OD) | 0.734 | 0.777 | -0.0432 | -0.1126 to +0.0261 | 0.221 | 0.664 |
| GSE144127 | RF | MI mean rank | 0.734 | 0.732 | +0.0017 | -0.0691 to +0.0724 | 0.963 | 0.963 |
| GSE144127 | RF | Sweeney3 | 0.734 | 0.767 | -0.0335 | -0.1000 to +0.0331 | 0.325 | 0.664 |
| GSE144127 | RF | Zak16 | 0.734 | 0.832 | -0.0983 | -0.1531 to -0.0435 | 0.000 | 0.002 |
| GSE144127 | MLP | Berry86 | 0.853 | 0.784 | +0.0690 | +0.0265 to +0.1115 | 0.001 | 0.009 |
| GSE144127 | MLP | Combined8 | 0.853 | 0.857 | -0.0041 | -0.0086 to +0.0003 | 0.069 | 0.277 |
| GSE144127 | MLP | Kaforou (ATB vs OD) | 0.853 | 0.781 | +0.0714 | +0.0232 to +0.1197 | 0.004 | 0.019 |
| GSE144127 | MLP | MI mean rank | 0.853 | 0.836 | +0.0163 | -0.0109 to +0.0436 | 0.240 | 0.479 |
| GSE144127 | MLP | Sweeney3 | 0.853 | 0.824 | +0.0290 | -0.0086 to +0.0665 | 0.130 | 0.391 |
| GSE144127 | MLP | Zak16 | 0.853 | 0.865 | -0.0122 | -0.0342 to +0.0097 | 0.274 | 0.479 |
| GSE144127 | singscore | Berry86 | 0.861 | 0.740 | +0.1215 | +0.0640 to +0.1790 | 0.000 | 0.000 |
| GSE144127 | singscore | Combined8 | 0.861 | 0.860 | +0.0014 | -0.0110 to +0.0138 | 0.824 | 1.000 |
| GSE144127 | singscore | Kaforou (ATB vs OD) | 0.861 | 0.794 | +0.0667 | +0.0141 to +0.1193 | 0.013 | 0.052 |
| GSE144127 | singscore | MI mean rank | 0.861 | 0.746 | +0.1148 | +0.0661 to +0.1635 | 0.000 | 0.000 |
| GSE144127 | singscore | Sweeney3 | 0.861 | 0.853 | +0.0081 | -0.0177 to +0.0338 | 0.539 | 1.000 |
| GSE144127 | singscore | Zak16 | 0.861 | 0.863 | -0.0017 | -0.0190 to +0.0156 | 0.845 | 1.000 |
| GSE144127 | DirAvg | Berry86 | 0.855 | 0.839 | +0.0165 | -0.0096 to +0.0425 | 0.216 | 0.647 |
| GSE144127 | DirAvg | Combined8 | 0.855 | 0.858 | -0.0029 | -0.0146 to +0.0089 | 0.631 | 1.000 |
| GSE144127 | DirAvg | Kaforou (ATB vs OD) | 0.855 | 0.821 | +0.0341 | -0.0033 to +0.0714 | 0.074 | 0.297 |
| GSE144127 | DirAvg | MI mean rank | 0.855 | 0.825 | +0.0303 | +0.0005 to +0.0602 | 0.046 | 0.232 |
| GSE144127 | DirAvg | Sweeney3 | 0.855 | 0.808 | +0.0478 | +0.0050 to +0.0906 | 0.029 | 0.173 |
| GSE144127 | DirAvg | Zak16 | 0.855 | 0.859 | -0.0032 | -0.0200 to +0.0135 | 0.703 | 1.000 |
| GSE152218 | LR | Berry86 | 0.963 | 0.997 | -0.0344 | -0.0890 to +0.0202 | 0.217 | 1.000 |
| GSE152218 | LR | Combined8 | 0.963 | 0.960 | +0.0026 | -0.0257 to +0.0310 | 0.855 | 1.000 |
| GSE152218 | LR | Kaforou (ATB vs LTB) | 0.963 | 0.958 | +0.0053 | -0.0183 to +0.0289 | 0.660 | 1.000 |
| GSE152218 | LR | MI mean rank | 0.963 | 0.936 | +0.0265 | -0.0180 to +0.0709 | 0.244 | 1.000 |
| GSE152218 | LR | Sweeney3 | 0.963 | 0.960 | +0.0026 | -0.0211 to +0.0264 | 0.827 | 1.000 |
| GSE152218 | LR | Zak16 | 0.963 | 0.950 | +0.0132 | -0.0136 to +0.0400 | 0.333 | 1.000 |
| GSE152218 | RF | Berry86 | 0.927 | 0.984 | -0.0569 | -0.1344 to +0.0207 | 0.151 | 0.904 |
| GSE152218 | RF | Combined8 | 0.927 | 0.960 | -0.0331 | -0.0843 to +0.0181 | 0.206 | 1.000 |
| GSE152218 | RF | Kaforou (ATB vs LTB) | 0.927 | 0.955 | -0.0278 | -0.0754 to +0.0198 | 0.253 | 1.000 |
| GSE152218 | RF | MI mean rank | 0.927 | 0.914 | +0.0132 | -0.0697 to +0.0961 | 0.755 | 1.000 |
| GSE152218 | RF | Sweeney3 | 0.927 | 0.960 | -0.0331 | -0.0837 to +0.0176 | 0.201 | 1.000 |
| GSE152218 | RF | Zak16 | 0.927 | 0.911 | +0.0159 | -0.0643 to +0.0960 | 0.698 | 1.000 |
| GSE152218 | MLP | Berry86 | 0.960 | 1.000 | -0.0397 | -0.0953 to +0.0159 | 0.162 | 0.971 |
| GSE152218 | MLP | Combined8 | 0.960 | 0.955 | +0.0053 | -0.0048 to +0.0154 | 0.303 | 1.000 |
| GSE152218 | MLP | Kaforou (ATB vs LTB) | 0.960 | 0.960 | +0.0000 | -0.0226 to +0.0226 | 1.000 | 1.000 |
| GSE152218 | MLP | MI mean rank | 0.960 | 0.944 | +0.0159 | -0.0297 to +0.0614 | 0.495 | 1.000 |
| GSE152218 | MLP | Sweeney3 | 0.960 | 0.960 | +0.0000 | -0.0213 to +0.0213 | 1.000 | 1.000 |
| GSE152218 | MLP | Zak16 | 0.960 | 0.944 | +0.0159 | -0.0093 to +0.0410 | 0.216 | 1.000 |
| GSE152218 | singscore | Berry86 | 0.955 | 0.987 | -0.0317 | -0.0849 to +0.0214 | 0.242 | 1.000 |
| GSE152218 | singscore | Combined8 | 0.955 | 0.942 | +0.0132 | -0.0114 to +0.0379 | 0.293 | 1.000 |
| GSE152218 | singscore | Kaforou (ATB vs LTB) | 0.955 | 0.966 | -0.0106 | -0.0346 to +0.0134 | 0.387 | 1.000 |
| GSE152218 | singscore | MI mean rank | 0.955 | 0.939 | +0.0159 | -0.0356 to +0.0673 | 0.545 | 1.000 |
| GSE152218 | singscore | Sweeney3 | 0.955 | 0.976 | -0.0212 | -0.0546 to +0.0123 | 0.215 | 1.000 |
| GSE152218 | singscore | Zak16 | 0.955 | 0.915 | +0.0397 | -0.0014 to +0.0807 | 0.058 | 0.349 |
| GSE152218 | DirAvg | Berry86 | 0.963 | 0.989 | -0.0265 | -0.0697 to +0.0168 | 0.230 | 0.921 |
| GSE152218 | DirAvg | Combined8 | 0.963 | 0.947 | +0.0159 | -0.0027 to +0.0345 | 0.094 | 0.565 |
| GSE152218 | DirAvg | Kaforou (ATB vs LTB) | 0.963 | 0.963 | +0.0000 | -0.0213 to +0.0213 | 1.000 | 1.000 |
| GSE152218 | DirAvg | MI mean rank | 0.963 | 0.944 | +0.0185 | -0.0232 to +0.0603 | 0.385 | 1.000 |
| GSE152218 | DirAvg | Sweeney3 | 0.963 | 0.944 | +0.0185 | -0.0270 to +0.0640 | 0.425 | 1.000 |
| GSE152218 | DirAvg | Zak16 | 0.963 | 0.926 | +0.0370 | -0.0067 to +0.0807 | 0.097 | 0.565 |
