## Supplementary material for "Cross-cohort benchmarking and validation reveals task-dependent generalisability in tuberculosis blood transcriptomic classification": S9 Table. Subgroup analyses in the external cohorts (directional average)

**Subgroup analyses in the external cohorts (directional average).**

AUC of each panel within clinical and demographic subgroups of the validation cohorts. The active TB group is held constant and the comparison group is restricted. Sample sizes are given as active TB versus comparator. The GSE152218 female subgroup contains a single active TB case and no AUC is estimable.

| Cohort | Subgroup | n (ATB vs comparator) | Nat (final) | Combined8 | Berry86 | Kaforou (ATB vs OD) | Kaforou (ATB vs LTB) | Sweeney3 | Zak16 |
| --- | --- | --- | --- | --- | --- | --- | --- | --- | --- |
| GSE144127 | OD = ‘Other’ only | 79 vs 147 | 0.869 | 0.876 | 0.864 | 0.841 | n.e. | 0.833 | 0.874 |
| GSE144127 | OD = LRTI/chest infection/URTI | 79 vs 32 | 0.906 | 0.91 | 0.866 | 0.89 | n.e. | 0.867 | 0.903 |
| GSE144127 | OD = LTBI (treatment indicated) | 79 vs 16 | 0.87 | 0.877 | 0.88 | 0.827 | n.e. | 0.799 | 0.866 |
| GSE144127 | OD = cancer | 79 vs 24 | 0.899 | 0.88 | 0.828 | 0.882 | n.e. | 0.814 | 0.896 |
| GSE144127 | OD = pneumonia | 79 vs 55 | 0.941 | 0.925 | 0.9 | 0.875 | n.e. | 0.846 | 0.938 |
| GSE144127 | OD = sarcoidosis | 79 vs 33 | 0.577 | 0.596 | 0.597 | 0.549 | n.e. | 0.576 | 0.591 |
| GSE144127 | OD excluding LTBI | 79 vs 284 | 0.854 | 0.857 | 0.836 | 0.821 | n.e. | 0.808 | 0.858 |
| GSE144127 | ethnicity = Black | 16 vs 50 | 0.87 | 0.882 | 0.861 | 0.886 | n.e. | 0.876 | 0.879 |
| GSE144127 | ethnicity = Indian | 39 vs 148 | 0.868 | 0.867 | 0.831 | 0.824 | n.e. | 0.816 | 0.86 |
| GSE144127 | ethnicity = White | 15 vs 71 | 0.9 | 0.905 | 0.89 | 0.853 | n.e. | 0.785 | 0.893 |
| GSE144127 | sex = Female | 28 vs 128 | 0.844 | 0.856 | 0.834 | 0.809 | n.e. | 0.766 | 0.852 |
| GSE144127 | sex = Male | 51 vs 172 | 0.863 | 0.859 | 0.838 | 0.828 | n.e. | 0.824 | 0.861 |
| GSE152218 | ATB BMI<16 vs LTB BMI<16, males only | 13 vs 6 | 0.872 | 0.833 | 0.949 | n.e. | 0.885 | 0.731 | 0.808 |
| GSE152218 | LTB BMI<16 only (malnourished vs malnourished) | 14 vs 16 | 0.946 | 0.924 | 0.982 | n.e. | 0.946 | 0.906 | 0.893 |
| GSE152218 | LTB BMI>=18.5 only | 14 vs 11 | 0.987 | 0.98 | 1.0 | n.e. | 0.987 | 1.0 | 0.974 |
| GSE152218 | sex = male | 13 vs 10 | 0.915 | 0.885 | 0.969 | n.e. | 0.931 | 0.838 | 0.877 |
