## Supplementary material for "Cross-cohort benchmarking and validation reveals task-dependent generalisability in tuberculosis blood transcriptomic classification": S10 Table. Direction preservation and leakage canary in the external cohorts

**Direction preservation and leakage canary in the external cohorts.**

Upper block: for every gene of the locked panels, the direction of change learned in the discovery cohorts, the per-cohort discovery AUC, and the AUC of that single gene in the validation cohort. Lower block: AUC when the validation expression matrix is replaced by noise, with labels unchanged.

| Panel | Gene | Direction in discovery | Discovery AUC per cohort | Validation AUC | Direction preserved |
| --- | --- | --- | --- | --- | --- |
| Nat (final) | APOL2 | up in ATB | GSE37250 0.787, GSE73408 0.752, GSE42834 0.807, GSE83456 0.796 | 0.792 | Yes |
| Nat (final) | WDFY1 | up in ATB | GSE37250 0.749, GSE73408 0.795, GSE42834 0.799, GSE83456 0.758 | 0.719 | Yes |
| Nat (final) | BATF2 | up in ATB | GSE37250 0.807, GSE73408 0.729, GSE42834 0.819, GSE83456 0.782 | 0.860 | Yes |
| Nat (final) | POLB | up in ATB | GSE37250 0.771, GSE73408 0.814, GSE42834 0.737, GSE83456 0.724 | 0.821 | Yes |
| Nat (final) | GBP2 | up in ATB | GSE37250 0.722, GSE73408 0.77, GSE42834 0.797, GSE83456 0.745 | 0.834 | Yes |
| MI mean rank | IFIT3 | up in ATB | GSE37250 0.721, GSE73408 0.708, GSE42834 0.791, GSE83456 0.678 | 0.795 | Yes |
| MI mean rank | CSGALNACT2 | down in ATB | GSE37250 0.333, GSE73408 0.251, GSE42834 0.466, GSE83456 0.539 | 0.521 | No |
| MI mean rank | APOL2 | up in ATB | GSE37250 0.787, GSE73408 0.752, GSE42834 0.807, GSE83456 0.796 | 0.792 | Yes |
| MI mean rank | SERPING1 | up in ATB | GSE37250 0.775, GSE73408 0.696, GSE42834 0.825, GSE83456 0.777 | 0.845 | Yes |
| Nat (final) | ANKRD22 | up in ATB | GSE19491 0.929, GSE37250 0.941, GSE73408 0.935, GSE101705 0.929, GSE107994 0.998 | 0.944 | Yes |
| Nat (final) | FCGR1A | up in ATB | GSE19491 0.928, GSE37250 0.968, GSE73408 0.939, GSE101705 0.922, GSE107994 0.999 | 0.958 | Yes |
| Nat (final) | PSTPIP2 | up in ATB | GSE19491 0.929, GSE37250 0.915, GSE73408 0.957, GSE101705 0.922, GSE107994 0.996 | 0.952 | Yes |
| MI mean rank | POLB | up in ATB | GSE19491 0.904, GSE37250 0.908, GSE73408 0.937, GSE101705 0.98, GSE107994 0.987 | 0.899 | Yes |
| MI mean rank | PSTPIP2 | up in ATB | GSE19491 0.929, GSE37250 0.915, GSE73408 0.957, GSE101705 0.922, GSE107994 0.996 | 0.952 | Yes |
| MI mean rank | LHFPL2 | up in ATB | GSE19491 0.929, GSE37250 0.96, GSE73408 0.943, GSE101705 0.911, GSE107994 0.999 | 0.894 | Yes |

| Cohort | LR | RF | MLP | singscore | DirAvg |
| --- | --- | --- | --- | --- | --- |
| GSE144127 | 0.576 | 0.490 | 0.547 | 0.543 | 0.542 |
| GSE152218 | 0.473 | 0.444 | 0.468 | 0.447 | 0.452 |
